# Gut microbiota dysbiosis observed in tuberculosis patients resolves partially with anti-tuberculosis therapy

**DOI:** 10.1101/2023.06.14.23291387

**Authors:** Sukanya Sahu, Sandeep R. Kaushik, Shweta Chaudhary, Amit kumar Mahapatra, Rukuwe Kappa, Wetesho Kapfo, Sourav Saha, Ranjit Das, Anjan Das, Vinotsole Khamo, Ranjan Kumar Nanda

## Abstract

**Objective:** *Mycobacterium tuberculosis* (Mtb) primarily affects the lungs with involvement of other organs causing tuberculosis (TB) in humans. Since the lung-gut axis is bidirectional, and the gut microbiota contributes to metabolic and immune homeostasis, we looked at the gut microbiota and metabolites of TB patients and controls, and whether the perturbations, if any, resolve with anti-tuberculosis treatment.

**Methods:** In this multicentric case-control study, a total of 107 fecal samples belonging to drug naïve active tuberculosis (ATB) patients and controls (non-tuberculosis: NTB and healthy), were collected from two clinical sites in India. A group of drug-naïve ATB patients (n=10) from one site was followed-up and monitored at 2, 4, 6, and 8 months of their anti-tuberculosis treatment. The fecal microbiome and metabolome of these study participants were characterized by 300 bp pair end sequencing of the V3-V4 region of 16S rRNA gene and gas chromatography-time of flight-mass spectrometry (GC-TOF-MS) respectively to identify disease and treatment-specific variations, if any.

**Results:** **Drug naïve** ATB and NTB patients showed a significant reduction of gut microbial diversity with respect to age matched healthy controls in both the clinical sites. ATB patient’s had underrepresentation of gut commensals such as *Faecalibacterium prausnitzii, Prevotella copri* DSM 18205, *Coprococcus catus,* and overrepresentation of *Clostridium difficile* ATCC 9689 = DSM 1296. Longitudinally followed-up ATB patients showed elimination of *Alkalihalobacillus* with treatment initiation, whereas harmful taxa such as *Stenotrophomonas* and *Klebsiella pneumoniae* appeared in treatment-completed subjects. Interestingly, the fecal metabolites also showed group-specific differences, clustering ATB patients away from the controls irrespective of the study sites. Consistently, fecal 2-piperidinone abundance was higher in ATB patients compared to healthy controls. The fecal metabolome of longitudinally followed-up ATB patients showed a gradual shift towards healthy during the course of treatment completion.

**Conclusion:** Gut microbial dysbiosis observed in tuberculosis patients at case presentation is partially resolved with 6 months of treatment completion and also reflected in their metabolite level. The observed microbial and metabolite imbalance in these ATB patients could explain disease pathology which needs further exploration to exploit their translational potential for therapeutics development.

## INTRODUCTION

Tuberculosis (TB) is caused by *Mycobacterium tuberculosis* (Mtb) infection and still a major global health problem. In 2021, ∼10.6 million new TB cases with ∼1.6 million deaths associated with it were reported worldwide (WHO report, 2022). [1] About one-third of the world’s total population is latently infected with TB and ∼10% of them may develop active TB (ATB) in their lifetime. [2]

The Mtb pathogenesis and disease outcome depends on multiple host and environmental factors including the host gut microbiome. Recent technological advances in the field of genomics have allowed us to detect the presence of numerous difficult to culture microbes in biological matrices. The presence of Mtb in the lungs alter the sputum microbiome composition in TB patients. The gut microbiota plays a critical role in maintaining a healthy immune/metabolic system and in many systemic and respiratory disease conditions including TB, it shows alteration. [3] [4] [5] [6] The immunomodulatory functions of the gut microbiota and its metabolites may prove to be critical in identifying targets that could modulate the host response against TB, in terms of reducing progression from latency, mitigating disease severity, and lowering the incidence of drug resistance and co-infection. [7] [8]

The current knowledge on gut microbiota dysbiosis in TB is limited to murine models and patients from specific settings with limited information on the effect of long anti-tuberculosis therapy. Multicentric studies may address the role of dietary habits and environmental factors in shaping the composition of the gut microbiota. Also documented were the effects of antibiotic and non-antibiotic medications on the gut microbial composition. [9] [10] In the current study, we aimed to identify deregulated fecal microbiome and metabolome in drug-naïve pulmonary ATB patients with respect to healthy and non-tuberculosis (NTB) controls, along with longitudinally followed-up ATB patients completing 6 months of treatment from two different clinical sites in India.

## MATERIALS AND METHODS

### Subject recruitment and classification

In this multi-centric, cross-sectional study, subjects were recruited from Nagaland (site-I) and Tripura (site-II) following the approved protocols of Nagaland Hospital Authority, Kohima (NHAK); Agartala Government Medical College, Agartala (AGMC); and International Centre for Genetic Engineering and Biotechnology, New Delhi. Subjects reporting symptoms of 2 weeks or more of cough, fever, weight loss, and night sweat to the outpatient departments of NHAK and AGMC and heathy subjects from these sites were enrolled. After receiving written signed informed consent forms, sputum samples (days 1 and 2) and fecal samples were collected at the day of reporting. Sputum samples were subjected to Ziehl-Nielsen acid-fast bacilli (AFB) sputum microscopy, culture, and GeneXpert (Cepheid, USA) analyses following WHO guidelines in Thyrocare Technologies, Navi Mumbai, a National Accreditation Board for Testing and Calibration Laboratory. Subjects with positive microscopy and culture or GeneXpert test results were grouped as active tuberculosis patients (ATB), and with all negative test results were grouped as non-tuberculosis patients (NTB) **(Figure 1A)**. These NTB subjects were clinically diagnosed with other pulmonary disease conditions like asthma, chronic obstructive pulmonary diseases (COPD), lung cancer, pneumonia, or suffering from more than one complication. The ATB patients received anti-tuberculosis treatment with 2 months of intensive (Isoniazid, Rifampin, Pyrazinamide, Ethambutol; HRZE) and 4 months of continuation phase as per the drug dose recommended by Revised National TB Control Programme (RNTCP) guidelines. A subset of ATB patients from site-I was longitudinally followed up at 2, 4, 6, and 8 months of initiating treatment and fecal samples were collected **(Figure 1B)**. Fecal samples were stored at -80°C within 4 hours of collection for further processing.

**Figure 1.**
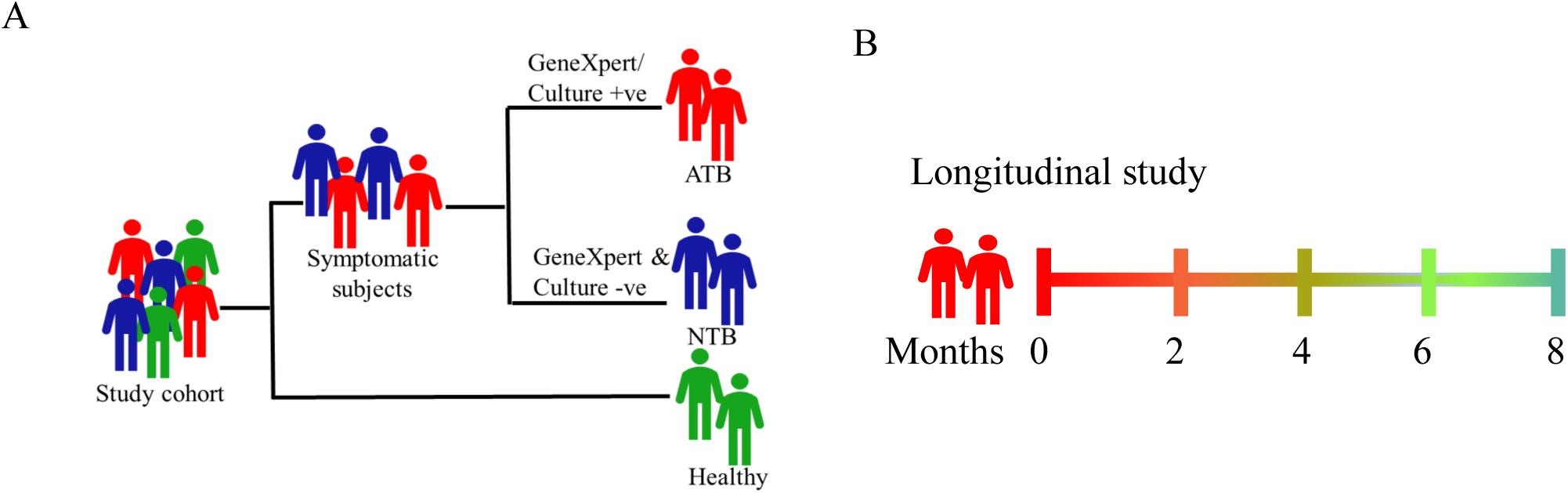
Strategy used for study subject recruitment and classification of A) case-control and B) longitudinally followed-up studies.

### DNA extraction and 16SrRNA gene amplicon sequencing

Genomic DNA from fecal samples were extracted following the earlier reported methods with minor modifications and stored at -20°C prior to analysis. [11] Quality of the extracted genomic DNA was monitored by agarose gel electrophoresis and gel eluted fractions were sent to a commercial laboratory (Macrogen, South Korea), for library preparation and sequencing. Briefly, the hypervariable region V3-V4 of bacterial 16S rRNA gene was amplified with the forward (Bakt_341F: CCTACGGGNGGCWGCAG) and the reverse primer (Bakt_805R: GACTACHVGGGTATCTAATCC). These 300bp PE V3-V4 amplicon libraries were sequenced using Illumina MiSeq. The detailed method of DNA extraction and library preparation is described in Supplemental Experimental Procedures. Raw sequence data generated in this study are assessable using the Sequence Read Archive under BioProject accession PRJNA972267.

### Sequence data processing, inferring gut microbiota composition, and statistical analyses

Quantitative Insights into Microbial Ecology (QIIME2 v.2022.2) tool was used for the 16S rRNA sequencing data analysis. [12] Non-singleton amplicon sequence variants (ASVs) were generated after quality filter, denoising, merging, and chimera removal using DADA2 plugin. Taxonomy was annotated with NCBI-Refseq database and classified at the level of phylum, class, order, family, genus, and species. Alpha diversity was characterized by Shannon and Faith PD indexes. The Kruskal-Wallis (KW) H test was used to assess the group specific differences. Beta diversity was described using unweighted and weighted UniFrac distance metrices and Permutational Analysis of Variance Analysis (PERMANOVA) was used to validate the differences between study groups. For the non-parametric test, Linear Discriminant Analysis with Effect Size (LEfSe) was used on the relatively normalized ASV table. [13] LefSe uses KW sum-rank test, pairwise Wilcoxon rank-sum test, and Linear discriminant analysis (LDA). Prediction of functional profiles from 16S rRNA datasets was conducted using Phylogenetic Investigation of Communities by Reconstruction of Unobserved States (PICRUSt2) software and the Kyoto Encyclopedia of Genes and Genomes (KEGG) database. [14] Metabolic pathway analysis was performed using PICRUSt2 to predict perturbed KEGG pathways in samples. These pathways were grouped into parent classes based on the KEGG BRITE hierarchy using R (v 4.2.1). Differential abundance analysis of KEGG pathways between groups was conducted using STAMP by KW H-test, followed by post-hoc Tukey-Kramer’s multiple comparison test, without any multiple test correction. [15]

### Faecal metabolite extraction and derivatization for GC-MS

To the lyophilized faecal samples (10.0 ± 0.3 mg; mean ± standard deviation, n=103), chilled methanol (1 ± 0.3 ml, −20 °C) along with lysine D4 (5 µl of 10 mg/ml) as a spike in standard was added. The sample mixture was vortexed (1 min), followed by ultrasonication at 20% amplitude (30 sec on/off cycle, total time 3 min), using QSonica Q500 sonicators (USA). Then the samples were vortexed, followed by centrifugation at 16,000 g for 15 min at 4 °C. A fraction of the supernatant (300 µl) was transferred to a micro centrifuge tube (MCT) and vacuum dried at 40 °C in SpeedVac vaccum concentrator (Labconco, USA). Aliquots of extracted metabolites from all the fecal samples (n=103) were pooled to prepare a quality control (QC). Derivatization of the fecal metabolites was carried out using earlier reported methods, and explained in detail in Supplemental Experimental Procedures. [16]

### GC-MS data acquisition

Using an automated multipurpose sample introduction system (MPS, Gerstel, Germany), the derivatized fecal metabolites (1 µl) were loaded into the column attached to GC-time of flight (TOF)-MS instrument (Pegasus 4D; Leco, USA) in splitless injection mode. Helium was used as a carrier gas with a constant flow rate of 0.5 ml/min for each run. Metabolite separation was carried out in an HP-5MS column (30 m× 0.25 mm × 0.25 µm; Agilent Technologies) with a temperature gradient from 60 °C to 300 °C at a ramp of 10 °C/min (60 to 220 °C) and 5 °C/min (220 to 300 °C). Hold times of 1 min and 5 min were kept at the start and end of the run, respectively. The injection port temperature was set at 250 °C throughout the analysis. Mass spectrometric data acquisition was carried out at -70 eV, and a mass range of 50-550 m/z was scanned with a rate of 50 spectra/sec. A 575 sec solvent delay was used and the source temperature was set at 220 °C. All GC-MS parameters were controlled using ChromaTOF software (V.4.50.8.0; Leco, USA) and data acquisition was completed.

### GC-MS data processing

Raw GC-MS data files (n=112) of all the QC (n=9) and study samples (n=103) were pre-processed and aligned using the "Statistical Compare" feature of ChromaTOF. For peak picking, the minimum peak width was set at 1.3 sec, and the signal-to-noise ratio (S/N) threshold was 75. For tentative molecular feature identification, mainlib, nist_ri, nist_msms2, nist_msms, and replib libraries from NIST were used. The maximum retention time difference was set at 0 sec, and for mass spectral match, the minimum spectral similarity was set at 600. Aligned peak information was exported to a ".csv" format, and molecules present in more than 50% of samples in at least one study group were included for the analysis. Manual data curation was carried out to align peaks and remove unsilylated molecules and silanes from the data matrix.

### Chemometric analysis

Metaboanalyst 5.0 was used for carrying out the univariate and multivariate statistical analyses using the identified molecular features as variables with their abundance. Missing values of variables were imputed with one-fifth of the minimum positive values. Normalization by reference feature, i.e. lysine D4 with auto-scaling, was carried out to obtain a near-normal distribution. A partial least square discriminant analysis (PLS-DA) model was built using metadata containing all the study samples and all qualifying molecular features. Analytes with a Variable Importance projection (VIP) score >1.0 from the PLS-DA were selected as important features to classify the study groups. For important feature selection, a fold change (FC) of at least 2.0 (log_2_FC>±1.0), with a student’s t-test with p-value<0.05 parameters were selected. Majority of the identified important features were validated by running commercial standards following similar pre-processing and GC-MS methods.

### Metabolomics—Microbiome data integration

The microbiome and metabolome data of the study subjects were integrated using the mixOmics R package (version 6.20.0). [17] Differential metabolites and microbial taxa at genus level identified in ATB, NTB, and healthy groups were correlated. The mixOmics regularised canonical correlation analysis (rCCA) function was used to generate clustered heatmaps. Euclidean distance was used to cluster the microbiome data in the rows and the metabolome data in the columns.

### Patient and public involvement

Patients or the public were not involved in the design, conduct, reporting or dissemination plans of this study.

## RESULTS

### Socio-demographic and sequencing characteristics of the study subjects

A total of 107 fecal samples of ATB, NTB and healthy groups were used for metagenomics (site-I: Nagaland, n = 45; site-II: Tripura, n = 31; followed-up, n = 31), and a subset of 94 samples was used for metabolomics (site-I: Nagaland, n = 40; site-II: Tripura, n = 30; followed-up, n = 24). Demographic details of the study subjects are presented in Supplementary Table S1 and S2. In site-I, the ATB subjects were younger than healthy controls whereas all study groups from site-II had similar age distribution (Supplementary Figure S1A, S1B).

### Microbial diversity differs in tuberculosis patients (ATB) from non-tuberculosis (NTB) and healthy controls

In total, 13,804,505 paired-end sequencing reads obtained from 107 fecal samples (an average of 1,27,894/sample) were processed (Supplementary Table S3, S4, S5). A significantly reduced alpha diversity was observed in ATB and NTB patients compared to healthy subjects, using Shannon and faith PD index, whereas ATB and NTB showed comparable diversity (Figure 2A, 2B). Furthermore, unweighted and weighted UniFrac analyses showed separate clustering of ATB and NTB from healthy subjects, which was statistically significant (Unweighted; ATB/healthy: p = 0.005, NTB/healthy: p = 0.002; Weighted; ATB/healthy p = 0.003, NTB/healthy p = 0.005) (Figure 2C, 2D), whereas comparable diversity was observed between ATB and NTB subjects (p = 0.303, p= 0.442, respectively). In site-II, a significantly lower Faith PD index was observed in ATB compared to the healthy controls (Figure 3A). Unweighted UniFrac analysis revealed a significant (p=0.003) difference in β diversity between ATB and healthy subjects, while it was comparable between ATB and NTB groups (Figure 3B).

**Figure 2.**
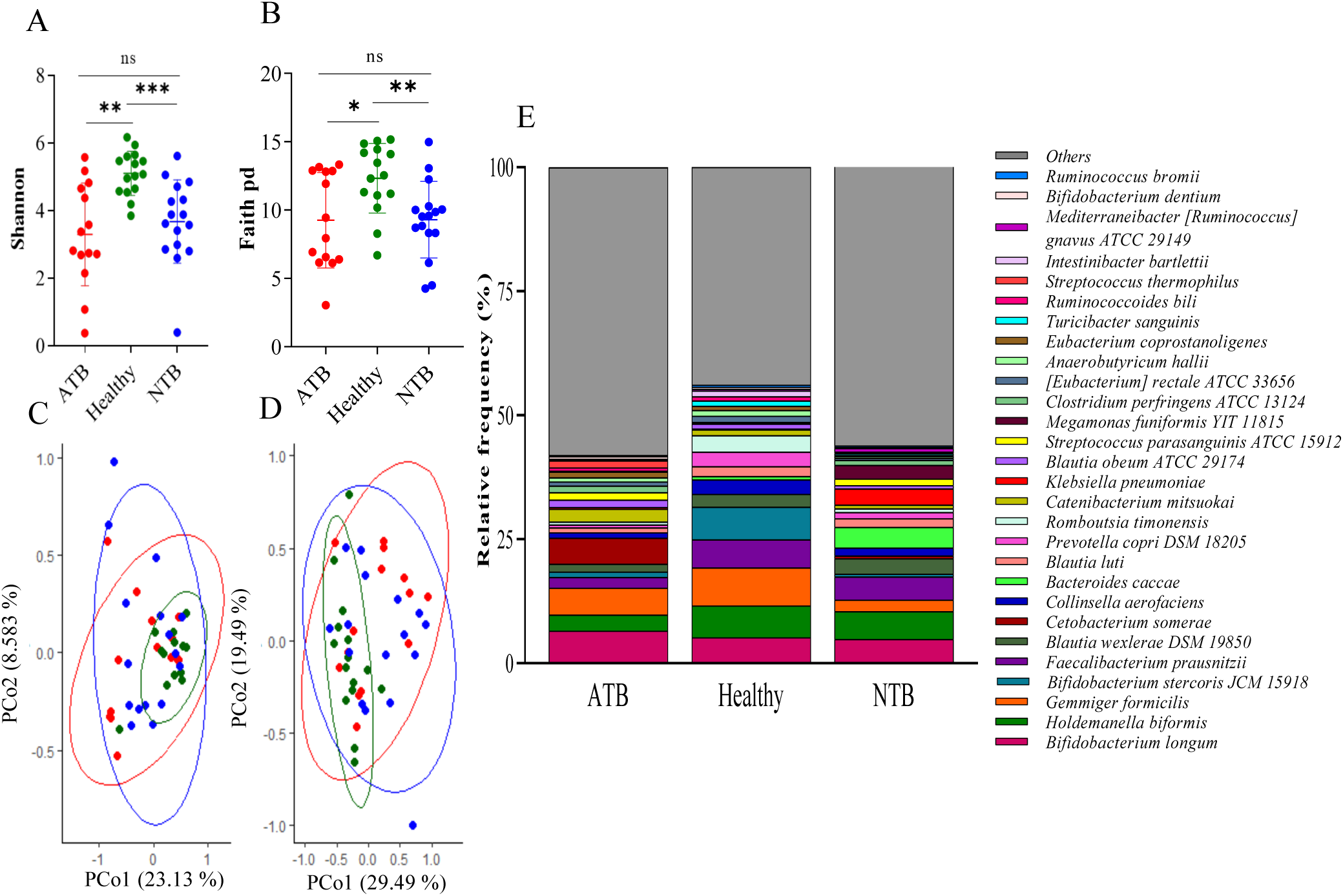
Gut microbial diversity analysis and taxonomic composition of drug naïve active tuberculosis patients (ATB), healthy and non-tuberculosis (NTB) controls from site-I. Alpha diversity comparisons based on A) Shannon and B) Faith PD index. Principal coordinate analysis (PCoA) of C) unweighted and D) weighted Unifrac distances. *: p<0.05; **: p<0.01; ***: p<0.001; ns: non-significant. E) Stacked bar plots representing the average percentage of top most taxa collapsed up to species level. Species that remained unclassified or with lesser abundance are grouped as others.

With respect to healthy controls, in ATB patients, relative abundance of *Collinsella*, *Holdemanella*, *Bifidobacterium*, and *Veillonella* genera was lower and *Escherichia* was higher in NTB in site-I (Figure S2A). A similar taxa abundance at genus level was observed between the study groups of site-II (Figure S2B). In site-I, *Gemmiger formicilis*, *Bifidobacterium stercosis* JCM 15918, *Prevotella copri* DSM 18205, and *Romboutsia timonensis* were relatively lower in ATB and NTB with respect to healthy, whereas *Faecalibacterium prausnitzii, Holdemanella biformis* were reduced in ATB. *Cetobacterium somerae* was higher in ATB; and *Bacteroides caccae*, *Klebsiella pneumoniae* were higher in NTB (Figure 2E). In site-II, *Erysipelatoclostridium ramosum*, *Mediterraneibacter [Ruminococcus] torques* were higher in ATB, whereas *Enterococcus faecalis* and *Phacaeicola dorei* were relatively low in ATB and NTB compared to healthy (Figure 3C).

**Figure 3.**
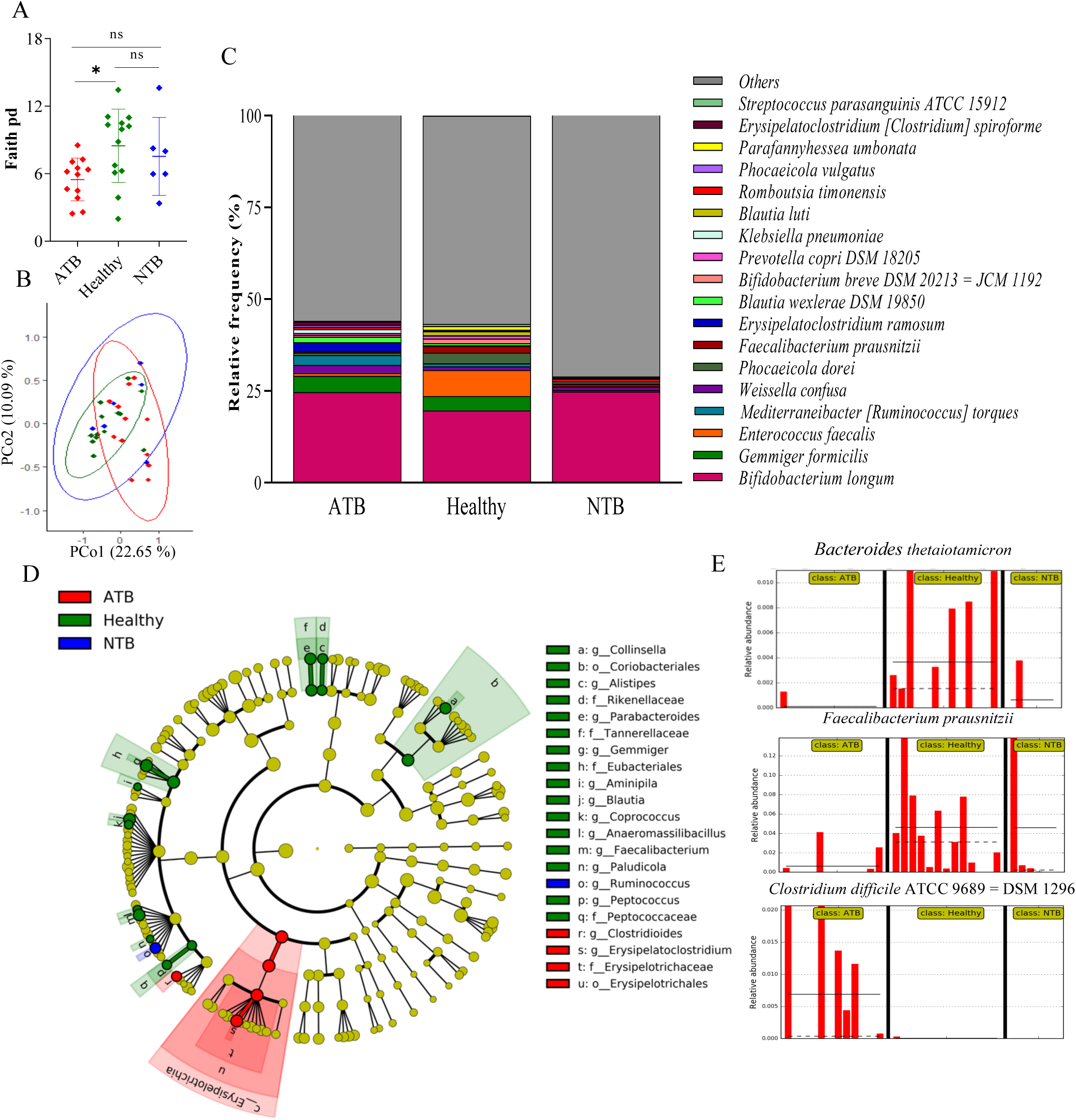
Gut microbial diversity analysis and taxonomic composition of drug naïve active tuberculosis patients (ATB), healthy and non-tuberculosis (NTB) controls from site-II. A) Alpha diversity comparisons based on Shannon index B) Principal coordinate analysis (PCoA) of unweighted Unifrac distance. *: p<0.05; **: p<0.01; ***: p<0.001; ns: non-significant. C) Stacked bar plots representing the average percentage of top most taxa collapsed up to species level. Species that remained unclassified or with lesser abundance are grouped as others. D) Cladogram representation of LEfSe analysis based on effect size (LDA score [log 10] threshold of 2. Differences among study groups were obtained by the Kruskal-Wallis test (α = 0.05) and Wilcoxon test (α = 0.05) with less strict parameter for multi-class analysis. E) Histogram of the relative abundances of identified species. The mean and median relative abundance are indicated with solid and dashed lines, respectively.

### Discrimination of ATB, healthy, and NTB subjects by gut bacterial markers

LEfSe analysis of study subjects from site-I revealed that *Dorea, Intestinibacter, Coprococcus, Prevotella, Collinsella,* and *Terrisporobacter* were depleted in ATB and NTB subjects with respect to healthy controls (Figure 4A, S3). *Alkalihalobacillus* was higher in ATB patients, whereas an increased abundance of *Klebsiella, Megamonas, Roseburia, Peptoniphilus,* and *Tyzzerella* was observed in NTB subjects (Figure 4A). A lower abundance of *Coprococcus catus, Prevotella copri* DSM 18205*, Lactobacillus rogosae, Collinsella bouchesdurhonensis, Collinsella aerofaciens, Senegalimassilia anaerobia* JC 110*, Intestinibacter bartletti,* and *Dorea longicatena* were observed in ATB and NTB subjects compared to healthy (Figure 4B, Supplementary Figure S4).

In site-II, *Erysipelatoclostridium* and *Clostridiodes* were higher in ATB, whereas *Faecalibacterium, Gemmiger, Blautia, Parabacteroides, Collinsella, Paludicola, Aminipila, Anaeromassilibacillus, Allistipes,* and *Peptococcus* were abundant including others in healthy (Figure 3D, S5). *Clostridium difficile* ATCC 9689 = DSM 1296 showed higher abundance in ATB subjects, whereas *Bacteroides thetaiotamicron, Faecalibacterium prausnitzii,* and *Parabacteroides merdae* were reduced in ATB compared to healthy (Figure 3E, S5).

### Prediction of functional potential

Variance analysis of KEGG metabolic pathways reflected differences in the functional genes in the microbiota of ATB and NTB from healthy groups of site-I (Figure 5A). Out of 39 active KEGG pathways identified at level 2, 12 pathways showed significant differences between ATB and healthy (p<0.05), and differences in 18 pathways were observed between healthy and NTB groups (Figure 5C). In site-II, study subjects showed similar clusters with a significant difference in the energy metabolism pathway between NTB and healthy (Figure 5B, 5D).

**Figure 5.**
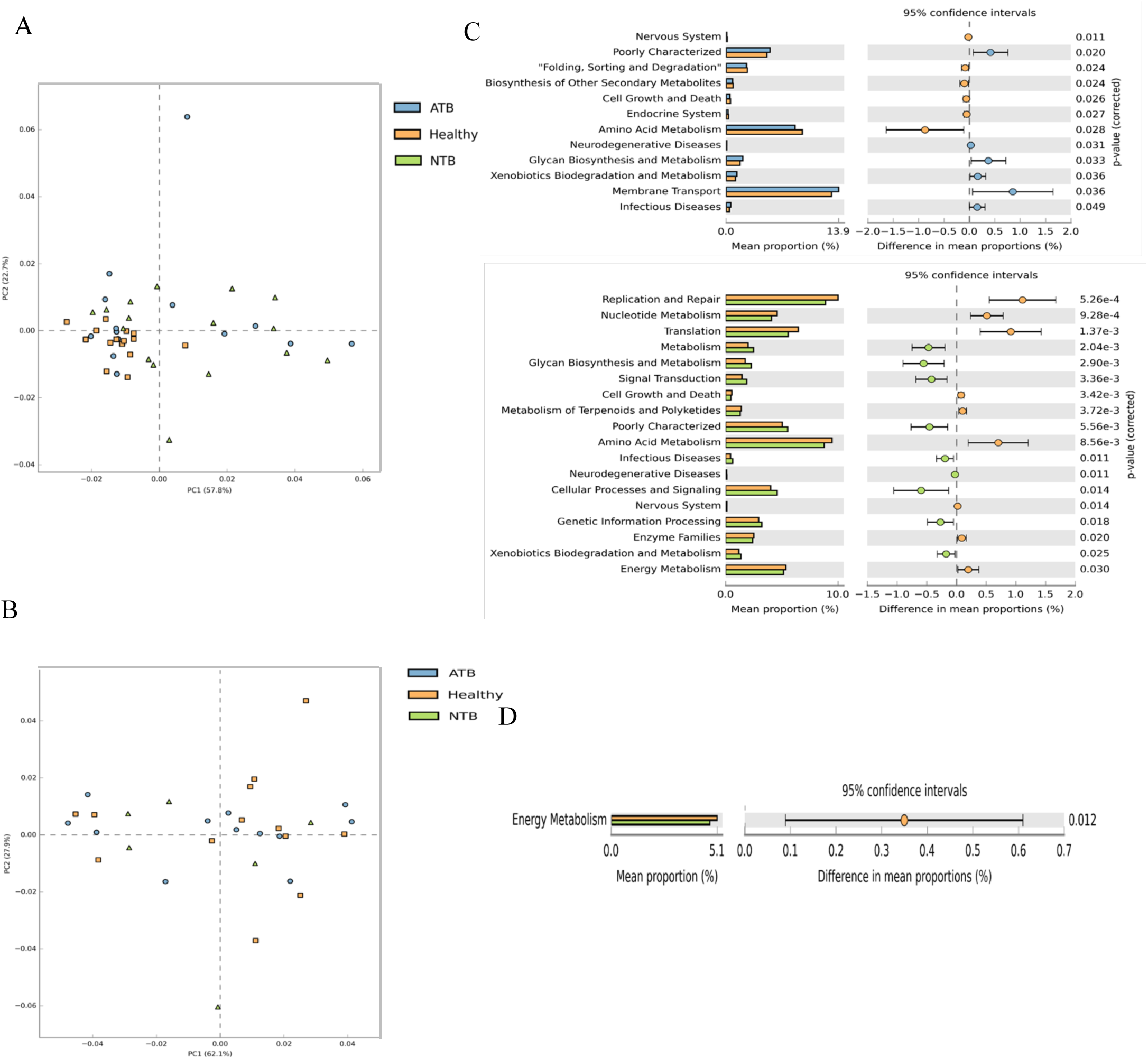
PICRUSt 2 analysis of study groups (active tuberculosis: ATB, healthy and non-tuberculosis: NTB) Statistical analysis was done using STAMP software. Principal Component Analysis plot of KEGG metabolic pathways in the second level, for multiple groups, applying Kruskal-Wallis H test, post-hoc Tukey-Kramer test for confidence interval method of study groups from A) site-I and B)Site-II. Extended error bar plot showed the mean proportions (%) of significantly different predicted functional categories at level 2 between study groups of C) site-I and D) site-II, applying Welch’s t-test, DP: Welch’s inverted test for confidence interval method. Variance analysis showed the abundance ratio of different functions in two groups of samples. The point showed the difference between proportions of functional abundance in the 95% confidence interval, and the value at the rightmost is the p-value. p<0.05 presents statistical significance.

### The fecal metabolome of ATB patients showed significant differences from controls

Global fecal metabolome analysis of ATB, healthy, and NTB groups, showed separate clusters between study groups from both the sites (Figure 6A, 6B). Set of differentially (FC ≥ 2, p ≤ 0.05) regulated metabolites between the study groups in site-I: ATB/healthy (low/high:11/13), ATB/NTB (low/high:11/3), NTB/healthy (low/high:8/14); and site-II: ATB/healthy (low/high:3/22), ATB/NTB (low/high: 6/2), NTB/healthy (low/high:1/22) were identified (Supplementary Figure S6A, S6C, S6E; S7A, S7C, S7E; Table S6-S11). A set of top 25 molecular features qualifying a VIP score > 1.0 and identified as important (Figure S6B, S6D, S6F; S7B, S7D, S7F). Elevated levels of 2-piperidinone were observed in ATB compared to healthy controls in both the study sites (Figure S6B, S7B).

**Figure 6.**
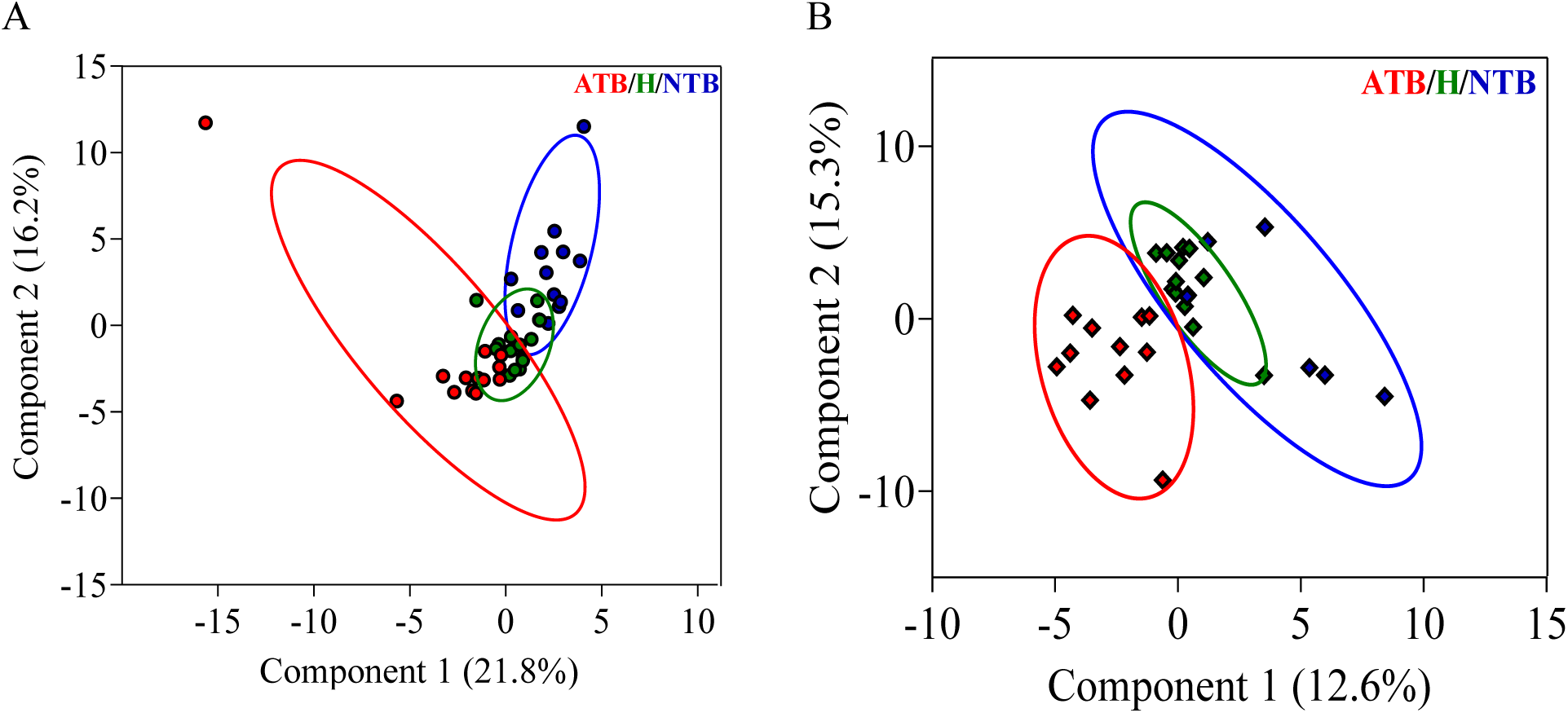
Fecal metabolites of active tuberculosis patients (ATB) showed significant variations from control (non-tuberculosis: NTB and healthy) subjects. PLS-DA score plot showing group specific clusters of fecal metabolites between ATB, Healthy and NTB Subjects of A) site-I and B) site-II. Coloured circles represent 95% confidence intervals. Coloured dots represent individual samples.

### Integrating microbiome and metabolome profile of ATB patients and controls

In site-I, dipterin, succinic acid, N-acetyl-L-Aspartic acid, and KGDS/1 showed a strong positive correlation with *Sarcina*. Similarly, *Romboustia*, *Senegalimassilia, Intestinibacter* showed a strong positive correlation (r^2^>0.8) with dihydrocinnamic acid and valeric acid in the healthy group. *Intestinibacter* negatively correlated with metronidazole, methylmalonic acid, DL-ornithine, and N-acetyl putrescine (Figure 7A). In site-II, *Erysipelatoclostridium*, enriched in ATB, showed a strong positive correlation (r^2^>0.8) with proline, 2-piperidinone, 4E-1-PDH-4(axial)-ol, 2,3-2H-Quinolin-2-one and Tris(hydroxymethyl)aminomethane, and was negatively correlated with tetradecanoic acid. Amino acids such as serine, L-ornithine, I-isoleucine, L-valine, DL-Phenylalanine, and I-aspartic acid showed a high negative correlation with *Faecalibacterium* abundance (Figure 7B). Other significant correlations between variables from both the study sites are detailed in the clustered image map (Figure 7A, 7B).

**Figure 7.**
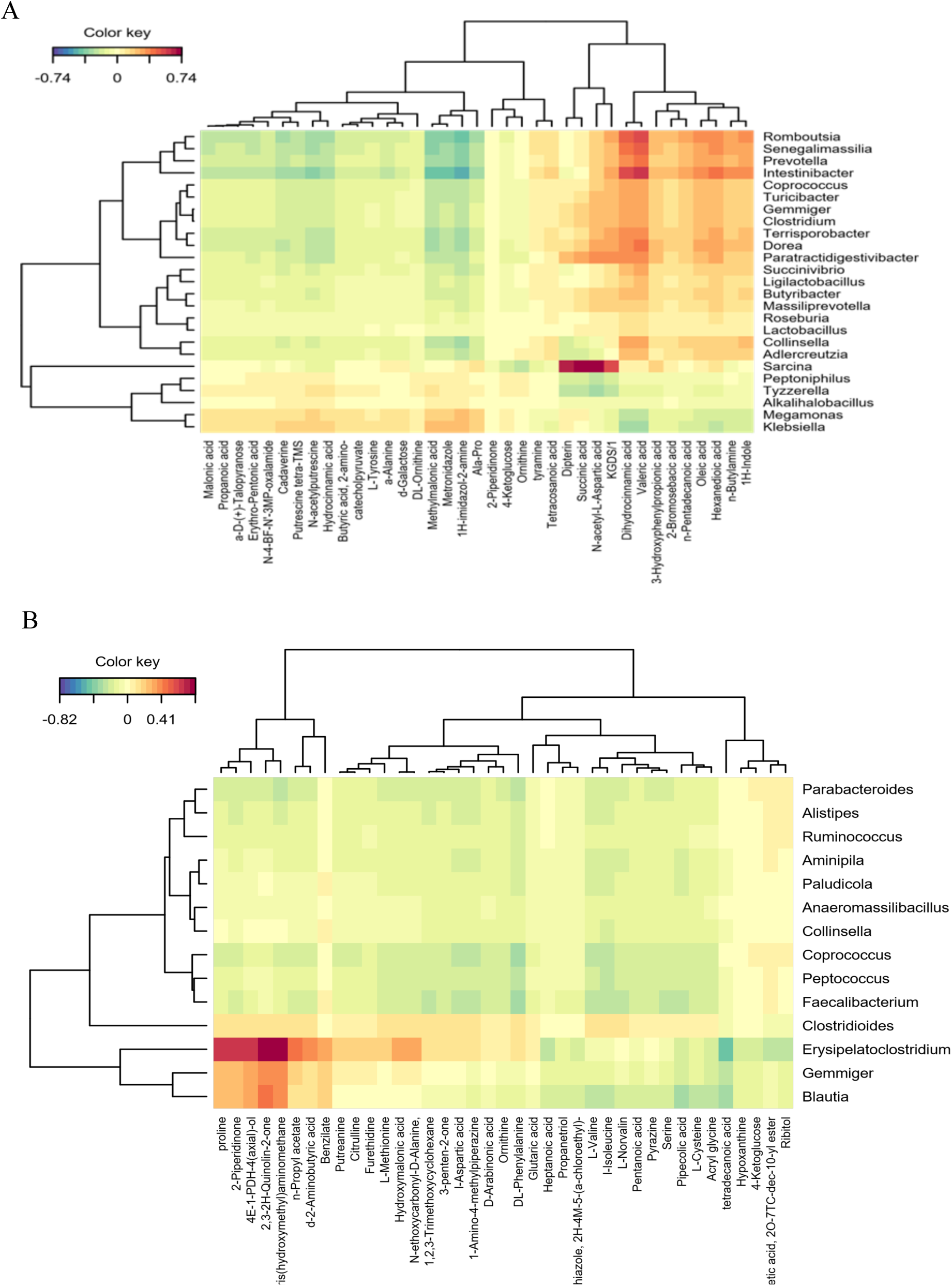
MixOmics analysis of study subjects from A) site-I and B) site-II. Clustered Image Map depicting correlation between differential fecal metabolic patterns and bacteria genera in ATB, healthy, and NTB subjects. Regularised canonical correlation coefficients between the level of reliable and markedly different fecal metabolic patterns and the abundance of the differentially enriched bacteria were calculated. Blue: negative correlation; Red: positive correlation.

### Tuberculosis patients receiving treatment revealed differential microbial and metabolite composition

Shannon index showed a time-dependent minor decrease in the α diversity of ATB subjects receiving anti-tuberculosis treatment (Figure 8A). β diversity analysis using unweighted UniFrac metric revealed comparable diversity of the study groups (Figure 8B) and PERMANOVA revealed significant differences between the 6 and 8 months of followed up subjects (p = 0.026). *Alkalihalobacillus* was abundant in drug-naïve ATB subjects, and significantly reduced upon treatment (Figure 8C). Whereas genera *Soonwooa*, *Stenotrophomonas, Diaphorobacter,* and *Agriterribacter* were observed in 8-month follow-up subjects, which was absent in drug-naïve ATB patients (Figure 8C, S8). *Ligilactobacillus ruminis*, *Diaphorabacter aerolatus, Actinomyces graevenitzii, Klebsiella pneumoniae* and *Agiterribacter humi* were observed in the 8-month follow-up subjects (Figure S8). *Soonwoa buanensis* was increased in 8-month followed-up subjects compared to drug-naïve ATB subjects (Figure S8). Multivariate analysis of the fecal metabolites of ATB patients at 0, 2, 4, 6, and 8 months of treatment completion and healthy subjects showed distinct clusters in PLS-DA analysis (Figure 8D). A set of 8 analytes, qualifying VIP score > 1.0, were selected as important features to group treatment specific variations in followed up cases (Figure S9). It was also observed that with treatment, the fecal metabolic profile of ATB subjects shifted towards the healthy group and may need longer time for complete overlapping (Figure 8D).

**Figure 8.**
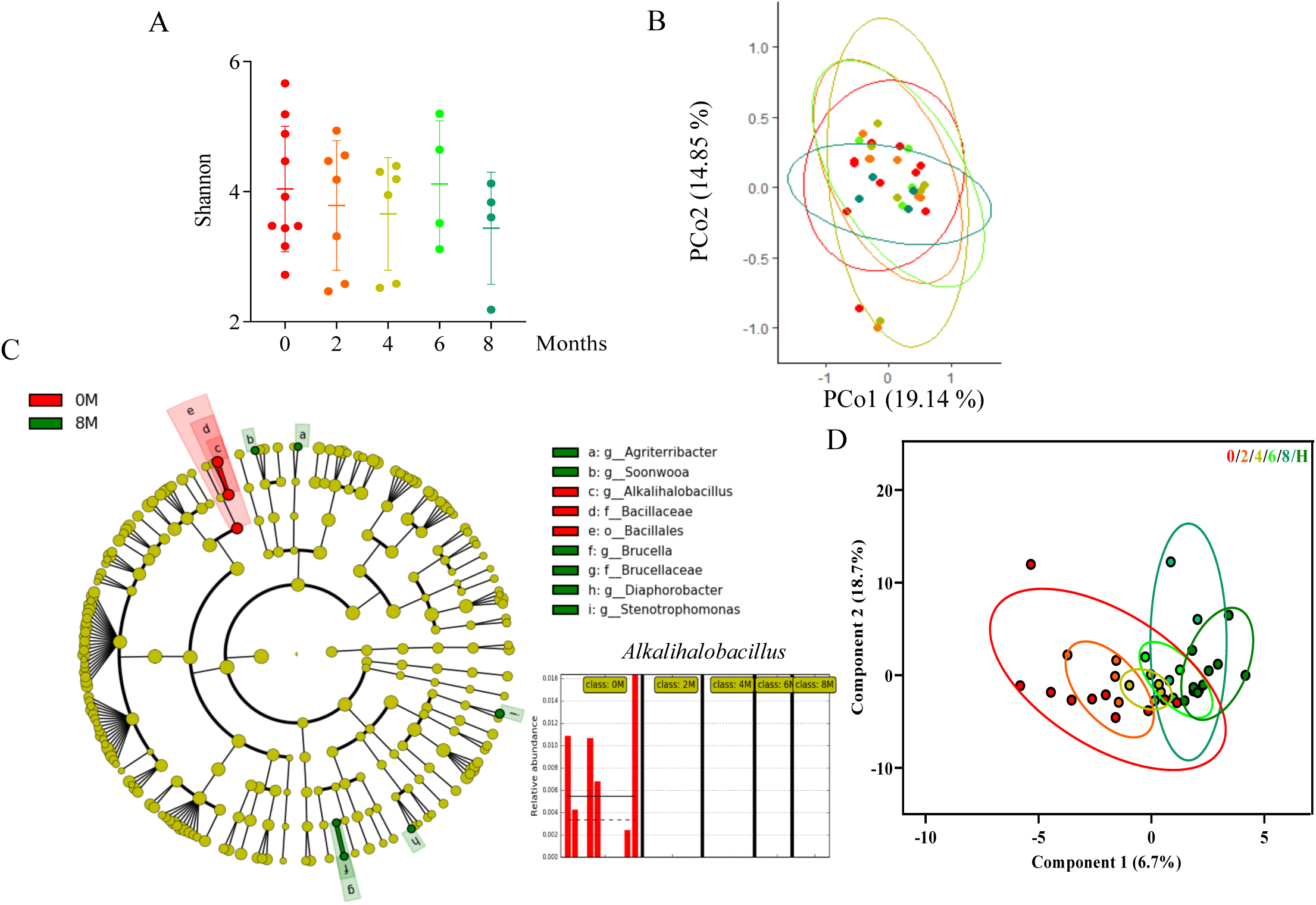
Fecal microbiota and metabolite study of longitudinally followed-up tuberculosis patients. A) Alpha diversity comparisons based on Shannon index B) Principal coordinate analysis (PCoA) of unweighted Unifrac distance. C) Cladogram representation of LEfSe analysis based on effect size (LDA score [log 10] threshold of 2. Differences among study groups were obtained by the Kruskal-Wallis test (α = 0.05) and Wilcoxon test (α = 0.05) with less strict parameter for multi-class analysis. Histogram of the relative abundances of Alkalihalobacillus. The mean and median relative abundance are indicated with solid and dashed lines, respectively. D) PLS-DA score plot for comparison of the global fecal metabolite profiles between 0,2,4,6,8 month and healthy subjects. Coloured circles represent 95% confidence intervals. Coloured dots represent individual samples.

## DISCUSSION

Since most of the studies have focussed on a single population, a multicentric study on the gut microbiota association with TB is the need of the hour. It is well established that gut dysbiosis in host negatively impacts the physiology and homeostatic processes. Earlier reports demonstrated significant loss of gut microbial diversity and taxonomic numbers in patients with pulmonary TB than those in healthy controls. [18] [19] In this study, we described the diversity, taxonomic composition of microbes, their predicted metabolic function, global metabolome differences in the gut microbiota of drug naive ATB patients compared to other disease (as NTB) or healthy controls and in a set of longitudinally followed up patients.

Our gut microbiome analysis of ATB and NTB patients revealed a significantly low α−diversity compared to healthy controls. The ATB and NTB groups had comparable microbial diversity whereas in site-I, NTB group presented with reduced diversity compared to healthy controls. β-diversity also demonstrated significant differences in the gut taxonomic composition, confirming intestinal microbiome changes in ATB. The richness in bacterial diversity is considered to have protective effects on metabolic and inflammatory diseases. [20] Therefore, the alterations in the intestinal flora of TB patients may be a signal for weakened anti-inflammatory ability.

Earlier reports demonstrated a significant loss of *Firmicutes* and an increased relative abundance of *Bacteroides* in the pulmonary TB group. [21] In contrast, we observed a comparable abundance of *Firmicutes* and *Bacteroides* in the ATB study groups. Proteobacteria were higher in NTB group. In addition, *Actinobacteria* is reduced in ATB compared to healthy and NTB, opposing earlier reports which may be partly be associated with the food habits, geographical origin of these subjects. [21]

At the genus level, higher abundance of SCFAs producing genera *Roseburia, Romboutsia, Butyribacter, Coprococcus, Lactobacillus* were observed in the healthy group. SCFAs are essential in maintaining a homeostatic environment, as they can induce either pro- or anti-inflammatory responses, depending on the signal transduction pathway. [22] *Roseburia* is reported to influence the production of SCFAs, namely, acetic, propionic and butyric acids and these metabolites protect the body from pathogens and inflammation. [23] [24] [25] In this study, Roseburia was reduced in the ATB patients, a result consistent with the study by Luo et al. [26] Thus, this underpin the likely involvement of SCFAs in TB pathogenesis. Decreased *Dorea, Roseburia* and *Ruminococcus* abundance was earlier reported in pulmonary TB patients. [19] Species level gut microbiota differences in TB patients have little been explored. Our study revealed the depletion of *Coprococcus catus* and *Collinsella aerofaciens* in ATB compared to healthy, which have been reported to produce propionate and butyrate, respectively.

The communication between the host and microbiome are partly contributed by metabolites, which profoundly influence the host physiology. Recent studies have uncovered that microbial metabolites play an important role in regulating the immune system. [27] By altering the production of butyric acid and propionic acid, the intestinal microbiota leads to impaired immune function in TB patients. [19] Reports revealed that the intestinal microbial metabolite indolepropionic acid (IPA) targets tryptophan to interfere with the metabolic activities of Mtb. [28] Fecal metabolome profile of ATB, healthy and NTB subjects from both the study sites showed distinct group specific clusters and a significant metabolic shift in ATB patients which corroborates earlier reports. [19] A set of 24 fecal metabolites showed significant alteration in ATB patients from both the sites with respect to healthy controls. One of them is 2-piperidinone, which showed significantly high abundance in ATB patients compared to healthy ones, and may have association with TB. ATB patients from site-I presented with reduction of valeric acid that corroborates an earlier report. [19] A set of 11 metabolites significantly altered in both ATB and NTB patients from healthy controls. Higher abundance of Hydrocinnamic acid, N-acetyl putrescine, Propanoic acid, methylmalonic acid and metronidazole and a significantly lower KGDS/1, N-acetyl-L-Aspartic acid, Hexanedioic acid, Dihydrocinnamic acid, succinic acid and valeric acid was observed between ATB and NTB subjects. Some drugs like metronidazole has been identified in the fecal samples and may be certain study subjects might have taken these drugs for resolving other issues.

Limited reports on the effect of anti-TB drugs on gut microbiota are available in literature and we monitored the gut microbiota in a set of longitudinally followed up drug-naïve TB patients. The clearance of *Alkalihalobacillus* along with appearance of certain opportunistic and harmful pathogens like *Stenotrophomonas* and *Klebsiella pneumoniae* indicates a partial restoration of gut microbiota in treatment completed TB patients. Further, metabolomics study revealed restoration of succinate, proline with treatment. Succinate, which is reported as the major metabolite of *Prevotella copri* could be the reason for the observed restoration of succinate with treatment. [29]

## CONCLUSION

Our findings provide direct first-hand evidence supporting gut microbial dysbiosis in drug naïve ATB patients with and without anti-tuberculosis treatment compared to healthy and other disease controls (NTB). However, the possibility that the gut microbiota dysbiosis precedes and contributes to the *M*. *tuberculosis* infection cannot be excluded. These findings need further validation in a larger population size from diverse patient population from developed, developing and underdeveloped countries to identify important microbes and their by-products, which may have translational potential in terms of pro- or post-biotics to manage TB patients for better therapeutic outcomes.

## Data Availability

All data produced are available online at https://www.ncbi.nlm.nih.gov/bioproject/PRJNA972267

## Acknowledgments

We would like to thank all the healthcare workers involved in the subject recruitment and sample collection from Naga Hospital Authority, Kohima and Agartala Government Medical College, Agartala. Dr Chaitali Nikam from Thyrocare Technologies Limited for helping us in the sputum culture tests that was used for subject classification.

## Contributors

RK, WK, SS, RD recruited study subjects and collated patients’ clinical data. SS and SRK performed all laboratory work including GeneXpert, DNA and metabolite extraction with assistance from AKM. SS analyzed and interpreted the sequencing data. SS, SRK and SC did the metabolite data acquisition and analysis. SS and RKN wrote the first draft of the manuscript and refined following suggestions of all co-authors.

## Patient consent for publication

Not required.

## Ethics approval

The study was approved by the institute review boards of Naga Hospital Authority, Kohima (NHAK) (NHAK/HLRC-008/2012 dated 17^th^ May 2017); Agartala Government Medical College, Agartala (AGMC) (protocolF.4[6-9]/AGMC/Academic/IEC Committee/2015/8965, dated 25 April 2018), and International Centre for Genetic Engineering and Biotechnology, New Delhi (ICGEB/IEC/2017/07). Written informed consent was obtained from all participants prior to collecting biological samples.

## Data availability statement

Data are available in a public, open access repository. https://www.ncbi.nlm.nih.gov/bioproject/PRJNA972267. Raw sequence data are available in the Sequence Read Archive (SRA) under BioProject accession PRJNA972267.

## References

1. World Health Organization. “Global tuberculosis programme” (2022). Report assessed on 11th June 2023.

2. Cohen, Adam, et al. "The global prevalence of latent tuberculosis: a systematic review and meta-analysis." European Respiratory Journal 54.3 (2019).

3. Schuijt, Tim J., et al. "The gut microbiota plays a protective role in the host defence against pneumococcal pneumonia." Gut 65.4 (2016): 575–583.

4. Gauguet, Stefanie, et al. "Intestinal microbiota of mice influences resistance to Staphylococcus aureus pneumonia." Infection and immunity 83.10 (2015): 4003–4014.

5. Ekbom, Anders, et al. "Increased risk of both ulcerative colitis and Crohn’s disease in a population suffering from COPD." Lung 186 (2008): 167–172.

6. Abrahamsson, T. R., et al. "Low gut microbiota diversity in early infancy precedes asthma at school age." Clinical & Experimental Allergy 44.6 (2014): 842–850.

7. Levy, Maayan, et al. "Dysbiosis and the immune system." Nature Reviews Immunology 17.4 (2017): 219–232.

8. Hu, Yongfeng, et al. "Gut microbiota associated with pulmonary tuberculosis and dysbiosis caused by anti-tuberculosis drugs." Journal of Infection 78.4 (2019): 317–322.

9. Patangia, Dhrati V., et al. "Impact of antibiotics on the human microbiome and consequences for host health." MicrobiologyOpen 11.1 (2022): e1260.

10. Weersma, Rinse K., Alexandra Zhernakova, and Jingyuan Fu. "Interaction between drugs and the gut microbiome." Gut 69.8 (2020): 1510–1519.

11. Bag, Satyabrata, et al. "An improved method for high quality metagenomics DNA extraction from human and environmental samples." Scientific reports 6.1 (2016): 26775.

12. QIIME 2 enables comprehensive end-to-end analysis of diverse microbiome data and comparative studies with publicly available data.

13. Segata, Nicola, et al. "Metagenomic biomarker discovery and explanation." Genome biology 12 (2011): 1–18.

14. Douglas, Gavin M., et al. "PICRUSt2 for prediction of metagenome functions." Nature biotechnology 38.6 (2020): 685–688.

15. Parks, Donovan H., et al. "STAMP: statistical analysis of taxonomic and functional profiles." Bioinformatics 30.21 (2014): 3123–3124.

16. Snijders, Antoine M., et al. "Influence of early life exposure, host genetics and diet on the mouse gut microbiome and metabolome." Nature microbiology 2.2 (2016): 1–8.

17. Rohart, Florian, et al. "mixOmics: An R package for ‘omics feature selection and multiple data integration." PLoS computational biology 13.11 (2017): e1005752.

18. Hu, Yongfei, et al. "The gut microbiome signatures discriminate healthy from pulmonary tuberculosis patients." Frontiers in cellular and infection microbiology 9 (2019): 90.

19. Wang, Shuting, et al. "Characteristic gut microbiota and metabolic changes in patients with pulmonary tuberculosis." Microbial biotechnology 15.1 (2022): 262–275.

20. Le Chatelier, Emmanuelle, et al. "Richness of human gut microbiome correlates with metabolic markers." Nature 500.7464 (2013): 541–546.

21. Wang, Yue, et al. "Alterations in the Gut Microbiome of Individuals With Tuberculosis of Different Disease States." Frontiers in Cellular and Infection Microbiology (2022): 328.

22. Eribo, Osagie A., et al. "The gut microbiome in tuberculosis susceptibility and treatment response: guilty or not guilty?." Cellular and Molecular Life Sciences 77 (2020): 1497–1509.

23. Estaki, Mehrbod, et al. "Cardiorespiratory fitness as a predictor of intestinal microbial diversity and distinct metagenomic functions." Microbiome 4 (2016): 1–13.

24. Rivière, Audrey, et al. "Bifidobacteria and butyrate-producing colon bacteria: importance and strategies for their stimulation in the human gut." Frontiers in microbiology 7 (2016): 979.

25. He, Jin, et al. "Short-chain fatty acids and their association with signalling pathways in inflammation, glucose and lipid metabolism." International journal of molecular sciences 21.17 (2020): 6356.

26. Luo, Mei, et al. "Alternation of gut microbiota in patients with pulmonary tuberculosis." Frontiers in physiology 8 (2017): 822.

27. Kim, Chang H. "Immune regulation by microbiome metabolites." Immunology 154.2 (2018): 220–229.

28. Negatu, Dereje Abate, et al. "Gut microbiota metabolite indole propionic acid targets tryptophan biosynthesis in Mycobacterium tuberculosis." MBio 10.2 (2019): e02781–18.

29. Liu, Juan, et al. "Association Between Intestinal Prevotella copri Abundance and Glycemic Fluctuation in Patients with Brittle Diabetes." Diabetes, Metabolic Syndrome and Obesity (2023): 1613–1621.

