## Supplemental Table S1, S2, S3,S4,S5,S6,S7,S8,S9,S10,S11, and Supplemental Figure S1, S2, S3,S4,S5,S6,S7,S8,S9 and Supplemental Method for "Gut microbiota dysbiosis observed in tuberculosis patients resolves partially with anti-tuberculosis therapy"

Supplementary Tables, Figures, and Methods

**Table S1** Epidemiological details of the all the study subjects used in fecal metagenomics study. A/NTB: active/non-tuberculosis; M: months; NA: not available

| | | Case-control study | | | | | | Follow-up study | | | | |
|---|---|---|---|---|---|---|---|---|---|---|---|---|
| | | Site-I | | | Site-II | | | Site-I | | | | |
| Demographic details | Total | ATB | NTB | Healthy | ATB | NTB | Healthy | 0M | 2M | 4M | 6M | 8M |
| Population size | 107 | 14 | 16 | 15 | 12 | 6 | 13 | 10 | 7 | 6 | 4 | 4 |
| Mean age (range) in years | 43.2 (18-76) | 35.64 (20-60) | 40.56 (18-70) | 50.2 (25-68) | 45.5 (25-70) | 52.17 (42-61) | 46.7 (19-70) | 38.20 (20-76) | 45 (24-76) | 45 (27-76) | 32.25 (23-55) | 45 (24-56) |
| Gender (Female/Male) | 32/75 | 4/10 | 1/15 | 6/9 | 4/8 | -/6 | 3/10 | 4/6 | 5/2 | 3/3 | 2/2 | -/4 |
| Body mass index (kg/m$^2$) | | 21.93 | 23.94 | 25.24 | NA | NA | NA | 20.51 | NA | NA | NA | NA |
| Cough (y/n/na) | 60/18/29 | 14/-/- | 15/1/- | -/-/15 | 11/-/1 | 6/-/- | 1/4/8 | 10/-/- | -/5/2 | 3/2/1 | -/3/1 | -/3/1 |
| Expectoration (y/n/na) | 47/25/35 | 10/3/1 | 12/4/- | -/-/15 | 10/1/1 | 6/-/- | -/-/13 | 8/2/- | -/5/2 | 1/4/1 | -/3/1 | -/3/1 |
| Haemoptysis (y/n/na) | 20/58/29 | 5/9/- | 4/12/- | -/-/15 | 3/8/1 | 4/2/- | -/5/8 | 4/6/- | -/5/2 | -/5/1 | -/3/1 | -/3/1 |
| Chest pain (y/n/na) | 38/40/29 | 8/6/- | 9/7/- | -/-/15 | 10/1/1 | 2/4/- | -/5/8 | 9/1/- | -/5/2 | -/5/1 | -/3/1 | -/3/1 |
| Breathlessness (y/n/na) | 34/44/29 | 8/6/- | 9/7/- | -/-/15 | 8/3/1 | 2/4/- | -/5/8 | 7/3/- | -/5/2 | -/5/1 | -/3/1 | -/3/1 |
| Wheeze (y/n/na) | 28/50/29 | 8/6/- | 7/9/- | -/-/15 | 6/5/1 | -/6/- | -/5/8 | 7/3/- | -/5/2 | -/5/1 | -/3/1 | -/3/1 |
| Body temp. (>37 °C/normal/na) | 34/45/28 | 8/6/- | 6/10/- | -/-/15 | 11/1/- | 4/2/- | -/5/8 | 5/5/- | -/5/2 | -/5/1 | -/3/1 | -/3/1 |
| Smoking (y/n/na) | 16/23/68 | -/4/10 | 3/13/- | -/-/15 | 8/3/1 | 3/3/- | 2/-/11 | -/-/10 | -/-/7 | -/-/6 | -/-/4 | -/-/4 |
| Alcoholism (y/n/na) | 20/37/50 | 1/3/10 | 7/9/- | 5/10/- | 5/6/1 | 2/4/- | -/5/8 | -/-/10 | -/-/7 | -/-/6 | -/-/4 | -/-/4 |
| Sputum AFB Test (+ve/-ve/na) | 23/45/39 | 10/4/- | -/16/- | -/-/15 | 4/8/- | -/-/6 | -/-/13 | 9/1/- | -/5/2 | -/5/1 | -/3/1 | -/3/1 |
| GeneXpert test result (+ve/-ve/na) | 35/24/48 | 12/1/1 | -/16/- | -/-/15 | 11/1/- | -/6/- | -/-/13 | 8/-/2 | 4/-/3 | -/-/6 | -/-/4 | -/-/4 |
| MGIT Culture test (+ve/-ve/na) | 13/39/55 | 5/9/- | -/16/- | -/-/15 | 4/8/- | -/-/6 | -/-/13 | 4/4/2 | -/2/5 | -/-/6 | -/-/4 | -/-/4 |
| Rif resistance (+ve/-ve/na) | 8/27/72 | -/12/2 | -/-/16 | -/-/15 | 8/3/1 | -/-/6 | -/-/13 | -/8/2 | -/4/3 | -/-/6 | -/-/4 | -/-/4 |

**Table S2** Epidemiological details of study subjects used in fecal metabolomics. A/NTB: active/non-tuberculosis; M: months; NA: not available

| | | Case-control study | | | | | | Follow-up study | | | | |
|---|---|---|---|---|---|---|---|---|---|---|---|---|
| | | Site-I | | | Site-II | | | Site-I | | | | |
| Demographic details | Total | ATB | NTB | Healthy | ATB | NTB | Healthy | 0 M | 2 M | 4 M | 6 M | 8 M |
| Population size | 94 | 13 | 12 | 15 | 11 | 6 | 13 | 9 | 4 | 4 | 3 | 4 |
| Mean age (range) in years | 45.2 (18-76) | 36.2 (20-60) | 40.2 (18-63) | 50.2 (25-68) | 47.1 (25-70) | 52.2 (42-61) | 46.7 (19-70) | 35.67 (20-76) | 52.8 (24-76) | 50.25 (27-76) | 35.3 (24-55) | 45 (24-56) |
| Gender (Female/Male) | 25/69 | 4/9 | 1/11 | 6/9 | 4/7 | -/6 | 3/10 | 3/6 | 2/2 | 1/3 | 1/2 | -/4 |
| Body mass index (kg/m$^2$) | | 22.33 | 24.35 | NA | NA | NA | NA | 20.27 | NA | NA | NA | NA |
| Cough (y/n/na) | 52/16/26 | 13/-/- | 11/1/- | -/-/15 | 10/-/1 | 6/-/- | 1/4/8 | 9/-/- | -/4/- | 2/2/- | -/2/1 | -/3/1 |
| Expectoration (y/n/na) | 41/21/32 | 10/2/1 | 8/4/- | -/-/15 | 9/1/1 | 6/-/- | -/-/13 | 7/2/- | -/4/- | 1/3/- | -/2/1 | -/3/1 |
| Haemoptysis (y/n/na) | 20/48/26 | 5/8/- | 4/8/- | -/-/15 | 3/7/1 | 4/2/- | -/5/8 | 4/5/- | -/4/- | -/4/- | -/2/1 | -/3/1 |
| Chest pain (y/n/na) | 33/35/26 | 7/6/- | 7/5/- | -/-/15 | 9/1/1 | 2/4/- | -/5/8 | 8/1/- | -/4/- | -/4/- | -/2/1 | -/3/1 |
| Breathlessness (y/n/na) | 30/38/26 | 8/5/- | 7/5/- | -/-/15 | 7/3/1 | 2/4/- | -/5/8 | 6/3/- | -/4/- | -/4/- | -/2/1 | -/3/1 |
| Wheeze (y/n/na) | 24/44/26 | 8/5/- | 6/6/- | -/-/15 | 5/5/1 | -/6/- | -/5/8 | 5/4/- | -/4/- | -/4/- | -/2/1 | -/3/1 |
| Body temp. (>37 ºC/normal/na) | 31/38/25 | 8/5/- | 5/7/- | -/-/15 | 10/1/- | 4/2/- | -/5/8 | 4/5/- | -/4/- | -/4/- | -/2/1 | -/3/1 |
| Smoking (y/n/na) | 15/18/61 | -/3/10 | 3/9/- | -/-/15 | 7/3/1 | 3/3/- | 2/-/11 | -/-/9 | -/-/4 | -/-/4 | -/-/3 | -/-/4 |
| Alcoholism (y/n/na) | 12/24/58 | 1/2/10 | 5/7/- | -/-/15 | 4/6/1 | 2/4/- | -/5/8 | -/-/9 | -/-/4 | -/-/4 | -/-/3 | -/-/4 |
| Sputum AFB Test (+ve/-ve/na) | 22/36/23 | 10/3/- | -/12/- | -/-/15 | 4/7/- | -/-/6 | -/-/13 | 8/1/- | -/4/- | -/4/- | -/2/1 | -/3/1 |
| GeneXpert test result (+ve/-ve/na) | 33/20/41 | 11/1/1 | -/12/- | -/-/15 | 10/1/- | -/6/- | -/-/13 | 9/-/- | 3/-/1 | -/-/4 | -/-/3 | -/-/4 |
| MGIT Culture test (+ve/-ve/na) | 13/32/49 | 5/8/- | -/12/- | -/-/15 | 3/8/- | -/-/6 | -/-/13 | 4/3/2 | -/2/2 | -/-/4 | -/-/3 | -/-/4 |
| Rif resistance (+ve/-ve/na) | 8/23/63 | -/11/2 | -/-/12 | -/-/15 | 8/2/1 | -/-/6 | -/-/13 | -/8/1 | -/2/2 | -/-/4 | -/-/3 | -/-/4 |

**Table S3** DADA2 summary statistics table for fecal samples of ATB, NTB, and healthy subjects from site-I

| Subjects id | Input | Filtered | Filtered (%) | Denoised | Merged | Merged (%) | Non-chimeric | Non-chimeric (%) |
|---|---|---|---|---|---|---|---|---|
| A1 | 112758 | 79228 | 70.26 | 77646 | 73907 | 65.54 | 64496 | 57.2 |
| A11 | 156643 | 100736 | 64.31 | 99655 | 97062 | 61.96 | 83714 | 53.44 |
| A18 | 111758 | 64996 | 58.16 | 63664 | 60177 | 53.85 | 38354 | 34.32 |
| A2 | 120889 | 84307 | 69.74 | 83541 | 81221 | 67.19 | 69129 | 57.18 |
| A21 | 121746 | 86056 | 70.68 | 85900 | 85250 | 70.02 | 79713 | 65.47 |
| A22 | 110948 | 74693 | 67.32 | 71963 | 66639 | 60.06 | 50656 | 45.66 |
| A23 | 118916 | 83186 | 69.95 | 82289 | 79004 | 66.44 | 61618 | 51.82 |
| A24 | 132195 | 93811 | 70.96 | 93569 | 93067 | 70.4 | 91305 | 69.07 |
| A25 | 123711 | 82989 | 67.08 | 80692 | 76142 | 61.55 | 59343 | 47.97 |
| A27 | 136003 | 84811 | 62.36 | 82014 | 76223 | 56.05 | 54822 | 40.31 |
| A4 | 133774 | 92092 | 68.84 | 91450 | 88743 | 66.34 | 57172 | 42.74 |
| A5 | 124175 | 83941 | 67.6 | 80959 | 74112 | 59.68 | 56944 | 45.86 |
| A6 | 131797 | 90972 | 69.02 | 89012 | 84394 | 64.03 | 59134 | 44.87 |
| A8 | 126737 | 85347 | 67.34 | 84123 | 82137 | 64.81 | 69485 | 54.83 |
| H1 | 158247 | 86417 | 54.61 | 84870 | 80256 | 50.72 | 63029 | 39.83 |
| H10 | 140986 | 79940 | 56.7 | 77470 | 71227 | 50.52 | 54266 | 38.49 |
| H11 | 152763 | 96080 | 62.89 | 92123 | 84227 | 55.14 | 64911 | 42.49 |
| H12 | 135582 | 81474 | 60.09 | 77830 | 69301 | 51.11 | 45815 | 33.79 |
| H13 | 130412 | 73166 | 56.1 | 72418 | 69216 | 53.07 | 56435 | 43.27 |
| H14 | 144621 | 83871 | 57.99 | 81086 | 74945 | 51.82 | 58849 | 40.69 |
| H15 | 149448 | 85769 | 57.39 | 82584 | 76071 | 50.9 | 61022 | 40.83 |
| H2 | 153948 | 94010 | 61.07 | 90586 | 82343 | 53.49 | 58282 | 37.86 |
| H3 | 147012 | 89913 | 61.16 | 86959 | 80439 | 54.72 | 62069 | 42.22 |
| H4 | 114958 | 62003 | 53.94 | 60678 | 56284 | 48.96 | 44224 | 38.47 |
| H5 | 129370 | 77943 | 60.25 | 75643 | 70811 | 54.74 | 53803 | 41.59 |
| H6 | 146368 | 91023 | 62.19 | 87017 | 78168 | 53.41 | 57861 | 39.53 |
| H7 | 133658 | 85399 | 63.89 | 82850 | 75362 | 56.38 | 55856 | 41.79 |
| H8 | 130584 | 72877 | 55.81 | 70722 | 65578 | 50.22 | 47491 | 36.37 |
| H9 | 134410 | 79320 | 59.01 | 76542 | 69536 | 51.73 | 52770 | 39.26 |
| N1 | 119804 | 82054 | 68.49 | 80159 | 74917 | 62.53 | 57842 | 48.28 |
| N10 | 125944 | 93046 | 73.88 | 92912 | 92677 | 73.59 | 91922 | 72.99 |
| N11 | 120415 | 82125 | 68.2 | 80897 | 76926 | 63.88 | 62505 | 51.91 |
| N12 | 121383 | 82589 | 68.04 | 81634 | 78949 | 65.04 | 67118 | 55.29 |
| N13 | 115124 | 78318 | 68.03 | 74877 | 69169 | 60.08 | 56154 | 48.78 |
| N14 | 106069 | 66031 | 62.25 | 65179 | 63086 | 59.48 | 53941 | 50.85 |
| N15 | 131180 | 83418 | 63.59 | 82554 | 78925 | 60.17 | 63682 | 48.55 |
| N16 | 129139 | 80797 | 62.57 | 77649 | 70662 | 54.72 | 53470 | 41.4 |
| N2 | 126367 | 81501 | 64.5 | 79349 | 73337 | 58.03 | 56420 | 44.65 |
| N3 | 126675 | 87261 | 68.89 | 86847 | 85432 | 67.44 | 66118 | 52.19 |
| N4 | 116165 | 70339 | 60.55 | 67866 | 60940 | 52.46 | 49661 | 42.75 |
| N5 | 117123 | 76462 | 65.28 | 74956 | 71200 | 60.79 | 52144 | 44.52 |
| N6 | 109971 | 72719 | 66.13 | 71756 | 68380 | 62.18 | 53283 | 48.45 |
| N7 | 113941 | 78964 | 69.3 | 77562 | 73078 | 64.14 | 54272 | 47.63 |
| N8 | 145173 | 103796 | 71.5 | 102830 | 100292 | 69.08 | 83973 | 57.84 |
| N9 | 127391 | 83131 | 65.26 | 81430 | 77184 | 60.59 | 63368 | 49.74 |

**Table S4** DADA2 summary statistics table of ATB, NTB and, healthy subjects from site-II

| Sample-id | Input | Filtered | Filtered (%) | Denoised | Merged | Merged (%) | Non-chimeric | Non-chimeric (%) |
|---|---|---|---|---|---|---|---|---|
| A33 | 131153 | 85291 | 65.03 | 84265 | 80670 | 61.51 | 61939 | 47.23 |
| A34 | 114984 | 78202 | 68.01 | 78070 | 77856 | 67.71 | 76097 | 66.18 |
| A35 | 136555 | 95495 | 69.93 | 94499 | 92209 | 67.53 | 69186 | 50.67 |
| A36 | 135069 | 82828 | 61.32 | 81946 | 78110 | 57.83 | 62630 | 46.37 |
| A37 | 110909 | 81444 | 73.43 | 80513 | 77962 | 70.29 | 58424 | 52.68 |
| A38 | 130818 | 83301 | 63.68 | 83058 | 82418 | 63 | 80519 | 61.55 |
| A39 | 117128 | 77230 | 65.94 | 76475 | 72799 | 62.15 | 48785 | 41.65 |
| A40 | 112011 | 79174 | 70.68 | 78954 | 78164 | 69.78 | 71482 | 63.82 |
| A41 | 107967 | 76992 | 71.31 | 76752 | 75773 | 70.18 | 62257 | 57.66 |
| A42 | 115794 | 67435 | 58.24 | 67323 | 66945 | 57.81 | 65085 | 56.21 |
| A44 | 123830 | 87140 | 70.37 | 86191 | 84185 | 67.98 | 70586 | 57 |
| A45 | 118590 | 78399 | 66.11 | 77592 | 75958 | 64.05 | 67051 | 56.54 |
| H16 | 136091 | 80729 | 59.32 | 79133 | 75216 | 55.27 | 54451 | 40.01 |
| H17 | 160167 | 98193 | 61.31 | 95683 | 90408 | 56.45 | 69697 | 43.52 |
| H18 | 152826 | 100572 | 65.81 | 100132 | 99577 | 65.16 | 96388 | 63.07 |
| H19 | 137387 | 77723 | 56.57 | 75182 | 69252 | 50.41 | 41790 | 30.42 |
| H20 | 146790 | 88563 | 60.33 | 87591 | 85948 | 58.55 | 74700 | 50.89 |
| H21 | 140212 | 85183 | 60.75 | 82964 | 78983 | 56.33 | 64156 | 45.76 |
| H22 | 100598 | 55353 | 55.02 | 54927 | 54177 | 53.85 | 49504 | 49.21 |
| H23 | 141565 | 91702 | 64.78 | 89269 | 82875 | 58.54 | 60211 | 42.53 |
| H24 | 124155 | 81514 | 65.66 | 81413 | 80902 | 65.16 | 75121 | 60.51 |
| H25 | 138703 | 81669 | 58.88 | 79780 | 75217 | 54.23 | 56373 | 40.64 |
| H26 | 125942 | 76631 | 60.85 | 76006 | 74165 | 58.89 | 56120 | 44.56 |
| H27 | 113523 | 62924 | 55.43 | 62798 | 62462 | 55.02 | 58774 | 51.77 |
| H28 | 134344 | 73499 | 54.71 | 71426 | 67091 | 49.94 | 51027 | 37.98 |
| N17 | 130475 | 81005 | 62.08 | 79671 | 76325 | 58.5 | 62914 | 48.22 |
| N18 | 124349 | 79349 | 63.81 | 76665 | 72627 | 58.41 | 57851 | 46.52 |
| N19 | 113569 | 68210 | 60.06 | 67738 | 67072 | 59.06 | 57596 | 50.71 |
| N20 | 131080 | 79761 | 60.85 | 78854 | 76434 | 58.31 | 51586 | 39.35 |
| N21 | 145276 | 97614 | 67.19 | 97311 | 96142 | 66.18 | 82829 | 57.01 |
| N22 | 147903 | 85650 | 57.91 | 85432 | 84947 | 57.43 | 79881 | 54.01 |

**Table S5** DADA2 summary statistics table of metagenomics data obtained from the longitudinally followed-up tuberculosis patients

| Sample-id | Input | Filtered | Filtered (%) | Denoised | Merged | Merged (%) | Non-chimeric | Non-chimeric (%) |
|---|---|---|---|---|---|---|---|---|
| 02A_12 | 135345 | 82421 | 60.9 | 80526 | 76554 | 56.56 | 55769 | 41.21 |
| 02A_13 | 131136 | 85627 | 65.3 | 84504 | 81349 | 62.03 | 63389 | 48.34 |
| 02A_14 | 127672 | 91717 | 71.84 | 89183 | 84882 | 66.48 | 63287 | 49.57 |
| 02A_16 | 113279 | 79013 | 69.75 | 77642 | 73232 | 64.65 | 51101 | 45.11 |
| 02A_26 | 158840 | 92037 | 57.94 | 91678 | 90162 | 56.76 | 78760 | 49.58 |
| 02A_28 | 119065 | 85368 | 71.7 | 84708 | 82196 | 69.03 | 74037 | 62.18 |
| 02A_9 | 116555 | 80144 | 68.76 | 78145 | 73261 | 62.86 | 51037 | 43.79 |
| 04A_14 | 139950 | 99970 | 71.43 | 98149 | 93312 | 66.68 | 71737 | 51.26 |
| 04A_16 | 127139 | 78917 | 62.07 | 77378 | 73745 | 58 | 50619 | 39.81 |
| 04A_20 | 118348 | 77840 | 65.77 | 77017 | 74447 | 62.91 | 57882 | 48.91 |
| 04A_26 | 114647 | 74045 | 64.59 | 73580 | 72356 | 63.11 | 57523 | 50.17 |
| 04A_28 | 97656 | 68764 | 70.41 | 68148 | 66310 | 67.9 | 52771 | 54.04 |
| 04A_3 | 123908 | 86839 | 70.08 | 85570 | 83057 | 67.03 | 62151 | 50.16 |
| 06A_12 | 123856 | 83625 | 67.52 | 81273 | 76256 | 61.57 | 59309 | 47.89 |
| 06A_13 | 127080 | 87041 | 68.49 | 85948 | 83635 | 65.81 | 66122 | 52.03 |
| 06A_3 | 111343 | 74680 | 67.07 | 73431 | 71209 | 63.95 | 54724 | 49.15 |
| 06A_30 | 121131 | 77827 | 64.25 | 77085 | 74170 | 61.23 | 55445 | 45.77 |
| 08A_10 | 120608 | 87692 | 72.71 | 86632 | 84756 | 70.27 | 66635 | 55.25 |
| 08A_13 | 130191 | 88724 | 68.15 | 86532 | 82744 | 63.56 | 66013 | 50.7 |
| 08A_16 | 126473 | 88208 | 69.74 | 86358 | 82849 | 65.51 | 63727 | 50.39 |
| 08A_40 | 127765 | 90345 | 70.71 | 89637 | 88215 | 69.04 | 72282 | 56.57 |
| A_10 | 116816 | 87448 | 74.86 | 86765 | 84478 | 72.32 | 69818 | 59.77 |
| A_12 | 129590 | 89332 | 68.93 | 86273 | 80337 | 61.99 | 57205 | 44.14 |
| A_13 | 115359 | 74836 | 64.87 | 74169 | 72054 | 62.46 | 55205 | 47.85 |
| A_14 | 123572 | 86640 | 70.11 | 83226 | 78410 | 63.45 | 60082 | 48.62 |
| A_16 | 133105 | 93621 | 70.34 | 89451 | 82759 | 62.18 | 65833 | 49.46 |
| A_20 | 153189 | 107548 | 70.21 | 105915 | 101727 | 66.41 | 77337 | 50.48 |
| A_26 | 121363 | 84297 | 69.46 | 83456 | 81137 | 66.85 | 58508 | 48.21 |
| A_3 | 112020 | 74772 | 66.75 | 73551 | 70649 | 63.07 | 46479 | 41.49 |
| A_7 | 123362 | 83166 | 67.42 | 81201 | 77140 | 62.53 | 59895 | 48.55 |
| A_9 | 128263 | 81907 | 63.86 | 79568 | 75668 | 58.99 | 60333 | 47.04 |

**Table S6** Important fecal metabolites (n=24) identified in drug naïve tuberculosis patients (ATB) from healthy controls recruited from site-I. FC: fold change

| Name | FC | $\log_2$(FC: ATB/healthy) | Raw. pval | $-\log_{10}$(p) |
|---|---|---|---|---|
| Erythro-Pentonic acid | 152.19 | 7.25 | 0.008 | 2.08 |
| 1H-imidazole-2-amine | 41.50 | 5.38 | 0.003 | 2.57 |
| Hydrocinnamic acid | 36.24 | 5.18 | 0.003 | 2.58 |
| Metronidazole | 33.34 | 5.06 | 0.018 | 1.74 |
| Malonic acid | 30.46 | 4.93 | 0.021 | 1.67 |
| catecholpyruvate | 20.86 | 4.38 | 0.031 | 1.50 |
| N-acetylputrescine | 20.63 | 4.37 | 0.004 | 2.38 |
| 2-Piperidinone | 8.78 | 3.13 | 0.046 | 1.34 |
| Methylmalonic acid | 8.27 | 3.05 | 0.013 | 1.89 |
| Propanoic acid | 6.78 | 2.76 | 0.025 | 1.61 |
| a-D-(+)-Talopyranose | 6.17 | 2.63 | 0.023 | 1.65 |
| d-Galactose | 5.57 | 2.48 | 0.016 | 1.79 |
| Butyric acid, 2-amino- | 4.05 | 2.02 | 0.019 | 1.72 |
| Dihydrocinnamic acid | 0.42 | -1.26 | 0.043 | 1.37 |
| Valeric acid | 0.29 | -1.80 | 0.016 | 1.79 |
| N-acetyl-L-Aspartic acid | 0.24 | -2.04 | 0.023 | 1.63 |
| tyramine | 0.17 | -2.57 | 0.042 | 1.37 |
| Tetracosanoic acid | 0.16 | -2.62 | 0.007 | 2.16 |
| Succinic acid | 0.14 | -2.79 | 0.037 | 1.43 |
| Hexanedioic acid | 0.14 | -2.87 | 0.031 | 1.50 |
| n-Pentadecanoic acid | 0.09 | -3.54 | 7.64E-05 | 4.12 |
| 1H-Indole | 0.06 | -4.14 | 0.005 | 2.29 |
| KGDS/1 | 0.03 | -4.92 | 0.002 | 2.60 |
| Oleic acid | 0.02 | -5.37 | 0.037 | 1.44 |

**Table S7** Important fecal metabolites (n=14) identified in drug naïve active tuberculosis patients (ATB) with respect to non-tuberculosis (NTB) controls from site-I. FC: fold change

| Name | FC | $log_2$(FC:ATB/NTB) | raw.pval | $-log_{10}$(p) |
|---|---|---|---|---|
| Erythro-Pentonic acid | 202.82 | 7.66 | 0.018 | 1.74 |
| Malonic acid | 18.46 | 4.21 | 0.044 | 1.36 |
| 2-Bromosebacic acid | 7.61 | 2.93 | 0.002 | 2.76 |
| Putrescine tetra-TMS | 0.28 | -1.83 | 0.033 | 1.48 |
| Ornithine | 0.28 | -1.84 | 0.007 | 2.17 |
| Dipterin | 0.22 | -2.16 | 0.011 | 1.97 |
| Oleic acid | 0.18 | -2.51 | 0.013 | 1.90 |
| 4-Ketoglucose | 0.16 | -2.68 | 0.008 | 2.07 |
| N-(4-Bromo-2-fluoro-phenyl)-N'-(3-morpholin-4-yl-propyl)-oxalamide | 0.09 | -3.42 | 0.049 | 1.31 |
| 1H-Indole | 0.08 | -3.56 | 0.040 | 1.40 |
| Tetracosanoic acid | 0.07 | -3.78 | 0.009 | 2.05 |
| Ala-Pro | 0.07 | -3.87 | 0.046 | 1.34 |
| 3-Hydroxyphenylpropionic acid | 0.05 | -4.33 | 0.037 | 1.44 |
| n-Pentadecanoic acid | 0.04 | -4.72 | 0.047 | 1.33 |

**Table S8** Important fecal metabolites (n=22) identified in non-tuberculosis (NTB) subjects with respect to healthy controls from site-I. FC: fold change

| Name | FC | $log_2$(FC:NTB/healthy) | raw.pval | -$log_{10}$(p) |
|---|---|---|---|---|
| Hydrocinnamic acid | 19.61 | 4.29 | 0.033 | 1.49 |
| N-acetylputrescine | 15.78 | 3.98 | 0.036 | 1.44 |
| Ala-Pro | 14.70 | 3.88 | 0.032 | 1.50 |
| N-(4-Bromo-2-fluoro-phenyl)-N'-(3-morpholin-4-yl-propyl)-oxalamide | 10.77 | 3.43 | 0.034 | 1.47 |
| Putrescine tetra-TMS | 9.90 | 3.31 | 0.003 | 2.49 |
| Metronidazole | 7.60 | 2.93 | 0.018 | 1.74 |
| Methylmalonic acid | 7.43 | 2.89 | 0.018 | 1.74 |
| 4-Ketoglucose | 6.44 | 2.69 | 0.005 | 2.32 |
| a-Alanine | 3.91 | 1.97 | 0.026 | 1.59 |
| Cadaverine | 3.81 | 1.93 | 0.001 | 2.92 |
| Ornithine | 3.59 | 1.85 | 0.004 | 2.41 |
| DL-Ornithine | 3.21 | 1.68 | 0.049 | 1.31 |
| Propanoic acid | 2.62 | 1.39 | 0.027 | 1.57 |
| L-Tyrosine | 2.17 | 1.12 | 0.031 | 1.50 |
| Valeric acid | 0.28 | -1.86 | 0.014 | 1.85 |
| Dihydrocinnamic acid | 0.24 | -2.05 | 0.003 | 2.58 |
| N-acetyl-L-Aspartic acid | 0.18 | -2.46 | 0.019 | 1.72 |
| 2-Bromosebacic acid | 0.17 | -2.54 | 0.025 | 1.60 |
| Succinic acid | 0.11 | -3.21 | 0.037 | 1.43 |
| Hexanedioic acid | 0.10 | -3.29 | 0.032 | 1.50 |
| n-Butylamine | 0.07 | -3.86 | 0.039 | 1.41 |
| KGDS/1 | 0.02 | -5.34 | 0.003 | 2.47 |

**Table S9** Important fecal metabolites (n= 25) identified in tuberculosis patients (ATB) from site-II study populations. FC: fold change

| **Name** | **FC** | **$\log_2$(FC:ATB/healthy)** | **raw.pval** | **$-\log_{10}$(p)** |
|---|---|---|---|---|
| proline | 84.41 | 6.40 | 0.023 | 1.64 |
| Furethidine | 68.32 | 6.09 | 0.049 | 1.31 |
| Putreanine | 47.81 | 5.58 | 0.027 | 1.57 |
| n-Propyl acetate | 36.65 | 5.20 | 0.008 | 2.10 |
| Hydroxymalonic acid | 35.56 | 5.15 | 0.012 | 1.91 |
| 2-Piperidinone | 31.27 | 4.97 | 0.009 | 2.04 |
| Citrulline | 26.86 | 4.75 | 0.017 | 1.77 |
| 1-Amino-4-methylpiperazine | 22.63 | 4.50 | 7.47E-04 | 3.13 |
| Acryl glycine | 19.74 | 4.30 | 0.035 | 1.45 |
| Glutaric acid | 19.64 | 4.30 | 0.044 | 1.36 |
| Pyrazine | 13.34 | 3.74 | 0.001 | 2.98 |
| N-ethoxycarbonyl-D-Alanine, | 10.67 | 3.42 | 0.049 | 1.31 |
| 1,2,3-Trimethoxycyclohexane | 10.16 | 3.34 | 0.011 | 1.96 |
| 3-penten-2-one | 9.93 | 3.31 | 4.29E-04 | 3.37 |
| Tris(hydroxymethyl)aminomethane | 7.66 | 2.94 | 4.45E-04 | 3.35 |
| D-Arabinonic acid | 7.43 | 2.89 | 0.036 | 1.45 |
| Ornithine | 6.43 | 2.69 | 0.021 | 1.67 |
| 2,3-2H-Quinolin-2-one | 6.29 | 2.65 | 0.025 | 1.60 |
| Pentanoic acid | 6.14 | 2.62 | 0.013 | 1.89 |
| 4(equat)Ethyl-1-n-propyl-trans-decahydroquinol-4(axial)-ol | 4.26 | 2.09 | 0.016 | 1.80 |
| L-Cysteine | 3.92 | 1.97 | 0.009 | 2.07 |
| Serine | 2.43 | 1.28 | 0.007 | 2.15 |
| tetradecanoic acid | 0.40 | -1.33 | 0.025 | 1.60 |
| Ribitol | 0.03 | -4.87 | 0.047 | 1.33 |
| Heptanoic acid | 0.03 | -5.21 | 0.030 | 1.52 |

**Table S10** Important fecal metabolites (n=23) identified in non-tuberculosis subjects (NTB) from site-II. FC: fold change

| Name | FC | $log_2$(FC: NTB/healthy) | raw. pval | $-log_{10}(p)$ |
|---|---|---|---|---|
| Acryl glycine | 38.78 | 5.28 | 0.021 | 1.67 |
| Propanetriol | 27.38 | 4.77 | 0.038 | 1.42 |
| 3-penten-2-one | 14.04 | 3.81 | 0.018 | 1.74 |
| proline | 13.58 | 3.76 | 0.003 | 2.52 |
| Citrulline | 13.14 | 3.72 | 0.014 | 1.86 |
| n-Propyl acetate | 10.94 | 3.45 | 0.023 | 1.64 |
| d-2-Aminobutyric acid | 9.16 | 3.20 | 0.013 | 1.89 |
| Ornithine | 8.80 | 3.14 | 0.014 | 1.85 |
| Pyrazine | 8.37 | 3.06 | 0.005 | 2.33 |
| N-ethoxycarbonyl-D-Alanine, | 6.98 | 2.80 | 0.043 | 1.37 |
| L-Cysteine | 6.32 | 2.66 | 0.015 | 1.83 |
| Hypoxanthine | 5.87 | 2.55 | 0.011 | 1.95 |
| Pipecolic acid | 5.82 | 2.54 | 0.011 | 1.94 |
| Serine | 5.77 | 2.53 | 0.019 | 1.72 |
| 1,2,3-Trimethoxycyclohexane | 4.88 | 2.29 | 0.007 | 2.14 |
| L-Valine | 4.70 | 2.23 | 0.009 | 2.03 |
| l-Isoleucine | 4.10 | 2.04 | 0.010 | 2.01 |
| Thiazole, 2-hydroxy-4-methyl-5-(a-chloroethyl)- | 4.06 | 2.02 | 0.007 | 2.18 |
| l-Aspartic acid | 4.04 | 2.01 | 0.010 | 2.02 |
| L-Methionine | 3.20 | 1.68 | 0.050 | 1.30 |
| Acetic acid, 2-oxa-7-thia-tricyclo[4.3.1.0(3,8)]dec-10-yl ester | 3.07 | 1.62 | 0.003 | 2.47 |
| DL-Phenylalanine | 2.64 | 1.40 | 0.036 | 1.45 |
| benzilate | 0.16 | -2.63 | 0.044 | 1.35 |

**Table S11** Important fecal metabolites (n=8) identified in active tuberculosis patients (ATB) compared to non-tuberculosis (NTB) subjects from site II. FC: fold change

| Name | FC | log2(FC:ATB/NTB) | raw. pval | -log10(p) |
|---|---|---|---|---|
| 1-Amino-4-methylpiperazine | 22.63 | 4.50 | 0.020 | 1.70 |
| Tris(hydroxymethyl)aminomethane | 7.66 | 2.94 | 0.015 | 1.83 |
| tetradecanoic acid | 0.46 | -1.11 | 0.036 | 1.45 |
| L-Norvalin | 0.38 | -1.41 | 0.027 | 1.56 |
| Acetic acid, 2-oxa-7-thia-tricyclo[4.3.1.0(3,8)]dec-10-yl ester/ Acetic acid, 2O-7TC-dec-10-yl ester | 0.33 | -1.62 | 0.007 | 2.14 |
| 4-Ketoglucose | 0.29 | -1.77 | 0.009 | 2.05 |
| Thiazole, 2-hydroxy-4-methyl-5-(a-chloroethyl)- | 0.25 | -2.02 | 0.013 | 1.90 |
| Hypoxanthine | 0.17 | -2.55 | 0.020 | 1.70 |

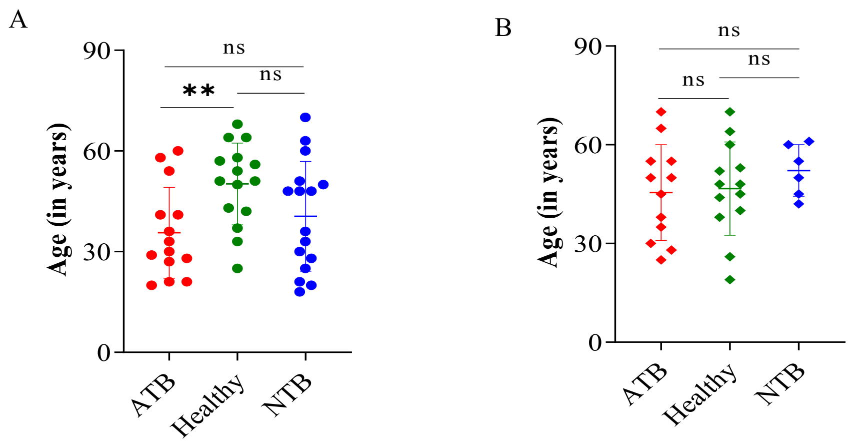


***Figure S1*** *Age distribution between study groups (active tuberculosis: ATB, non-tuberculosis: NTB and healthy subjects in A) site-I and B) site-II. The age data expressed as mean ± standard deviation (SD) and the difference between groups was evaluated by two-tailed Student's t test. **: p<0.01; ns: non-significant.*

A

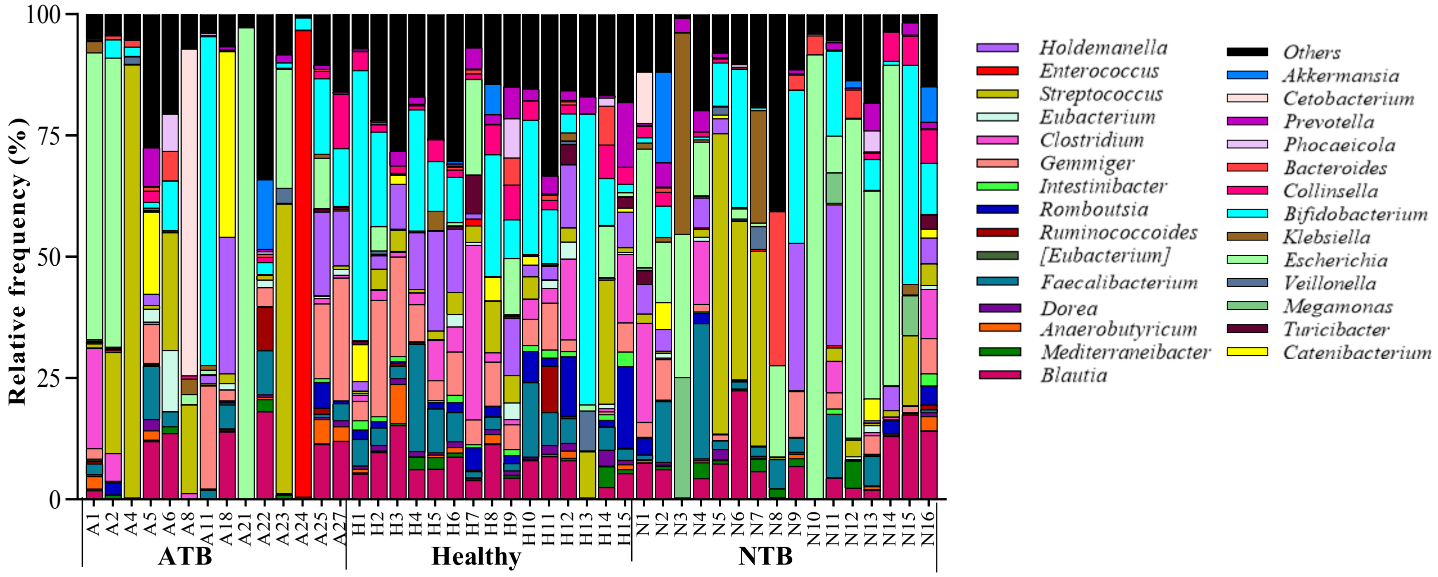


B

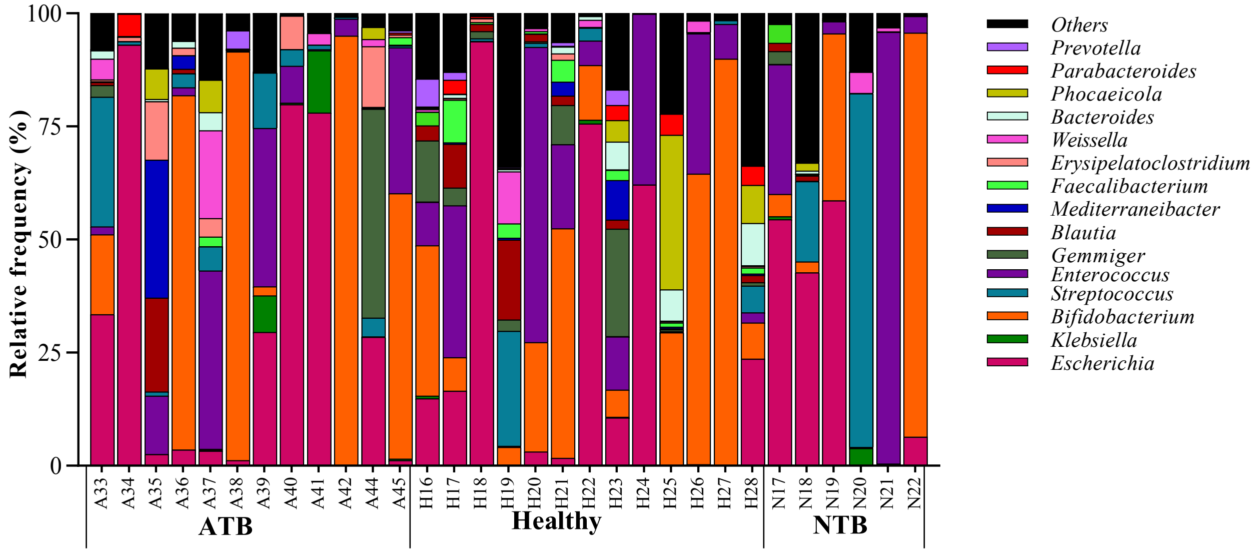


***Figure S2*** *Stacked bar plots showing the relative abundance of microbiota distribution in active tuberculosis patients (ATB), healthy and non-tuberculosis (NTB) subjects of A) site-I and B) site-II, with taxonomic features collapsed at the level of genus. Genus that remained unclassified or with lesser abundance are grouped as Others.*

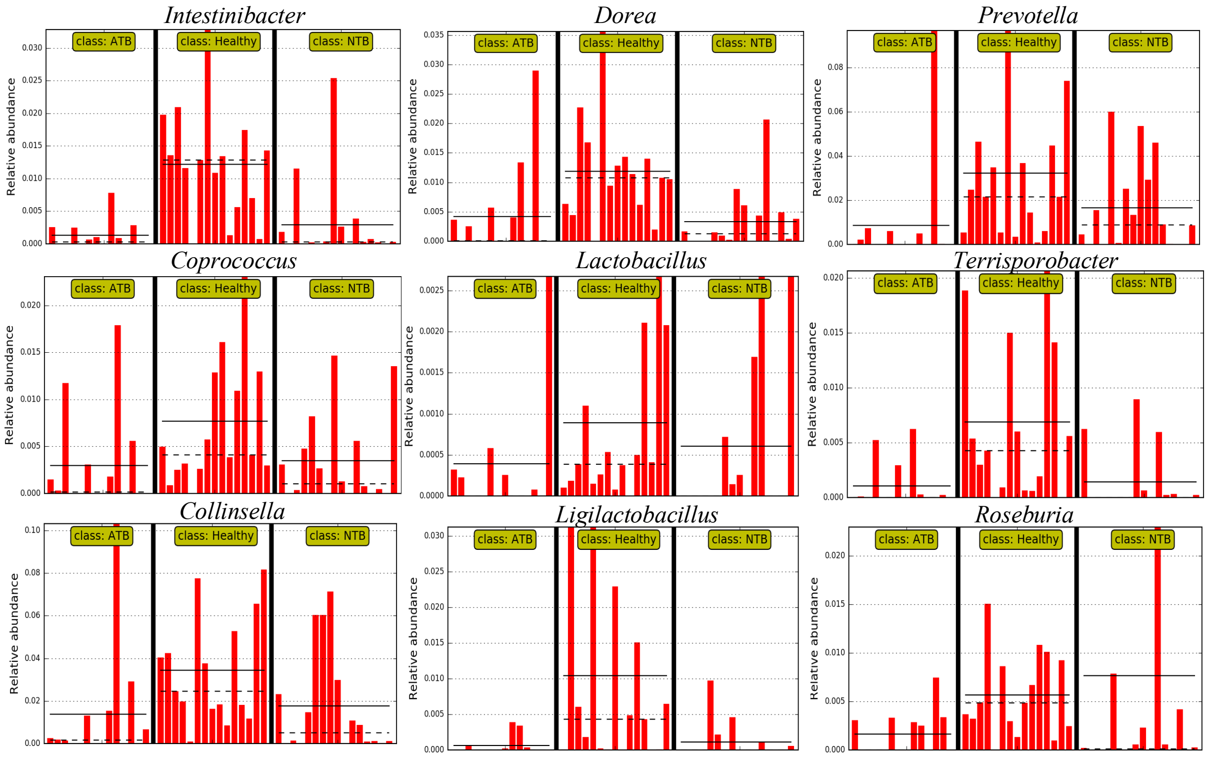


***Figure S3*** *LEfSe analysis of active tuberculosis patients (ATB), non-tuberculosis (NTB) and healthy subjects of site-I based on effect size (LDA score [log 10] threshold of 2. Differences among study groups were obtained by the Kruskal-Wallis test (α = 0.05) and Wilcoxon test (α = 0.05) with less strict parameter for multi-class analysis. The mean and median relative abundance of the genus are indicated with solid and dashed lines, respectively.*

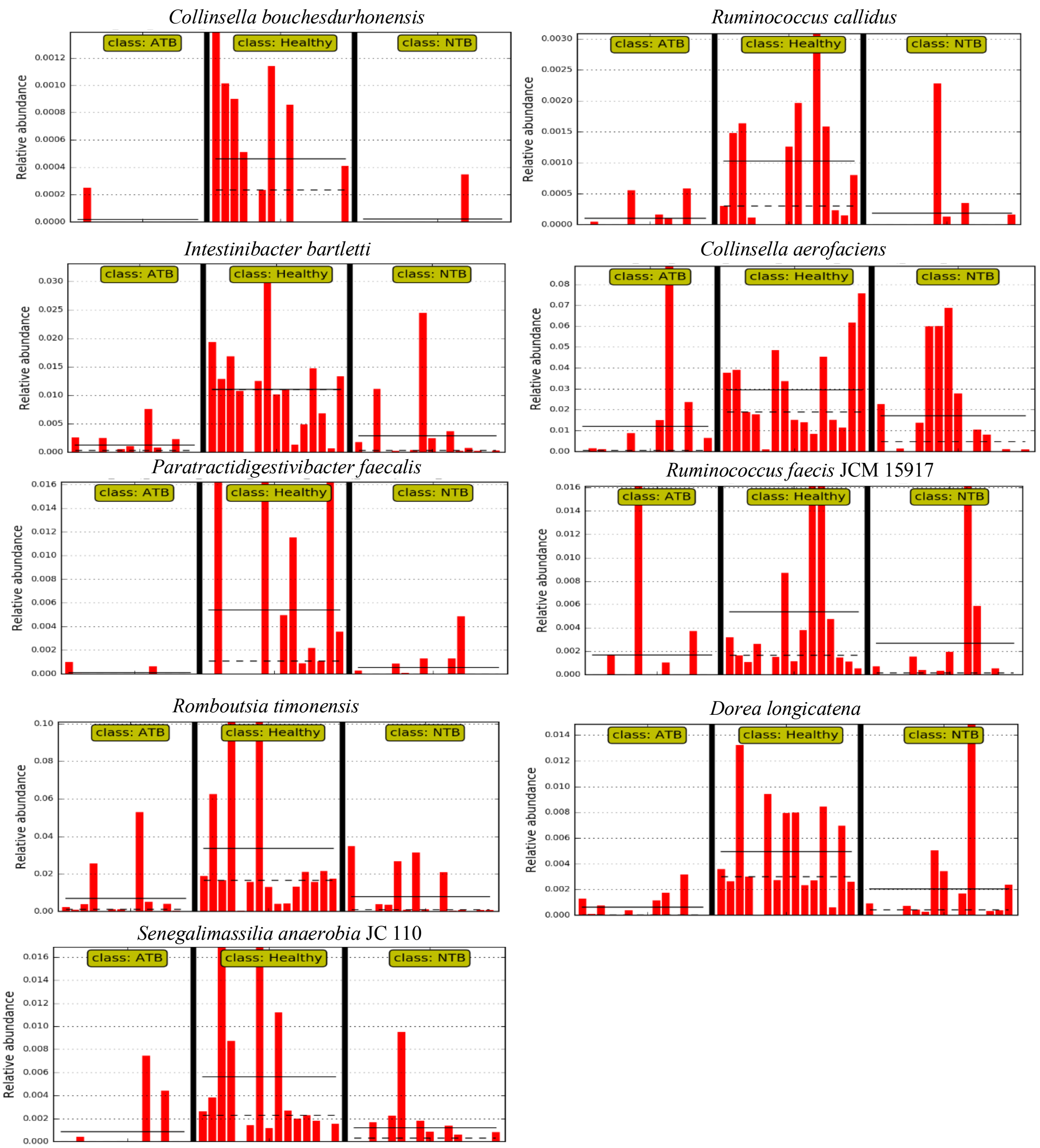


***Figure S4*** *Histogram showing the relative abundances of identified species in drug naïve tuberculosis patients (ATB), and controls (healthy and non-tuberculosis: NTB) of site-I by LEfSe analysis. The mean and median relative abundance of the genus are indicated with solid and dashed lines, respectively.*

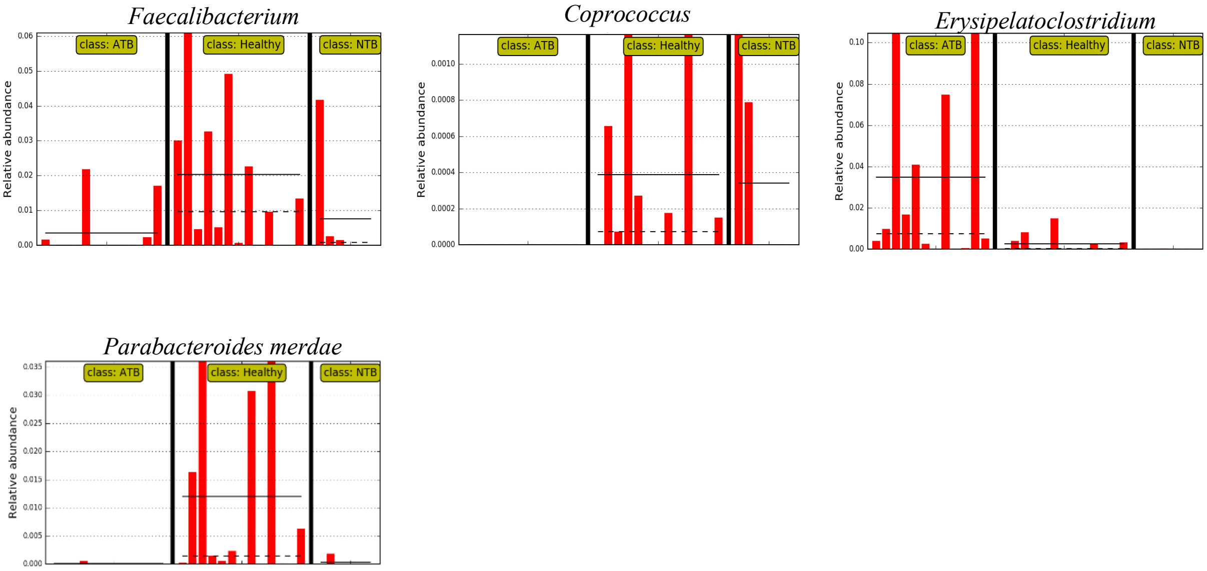


***Figure S5*** *Histogram showing the relative abundances of identified species in drug naïve tuberculosis patients (ATB), and controls (healthy and non-tuberculosis: NTB) of site-II by LEfSe analysis. The mean and median relative abundance of the genus and species are indicated with solid and dashed lines, respectively.*

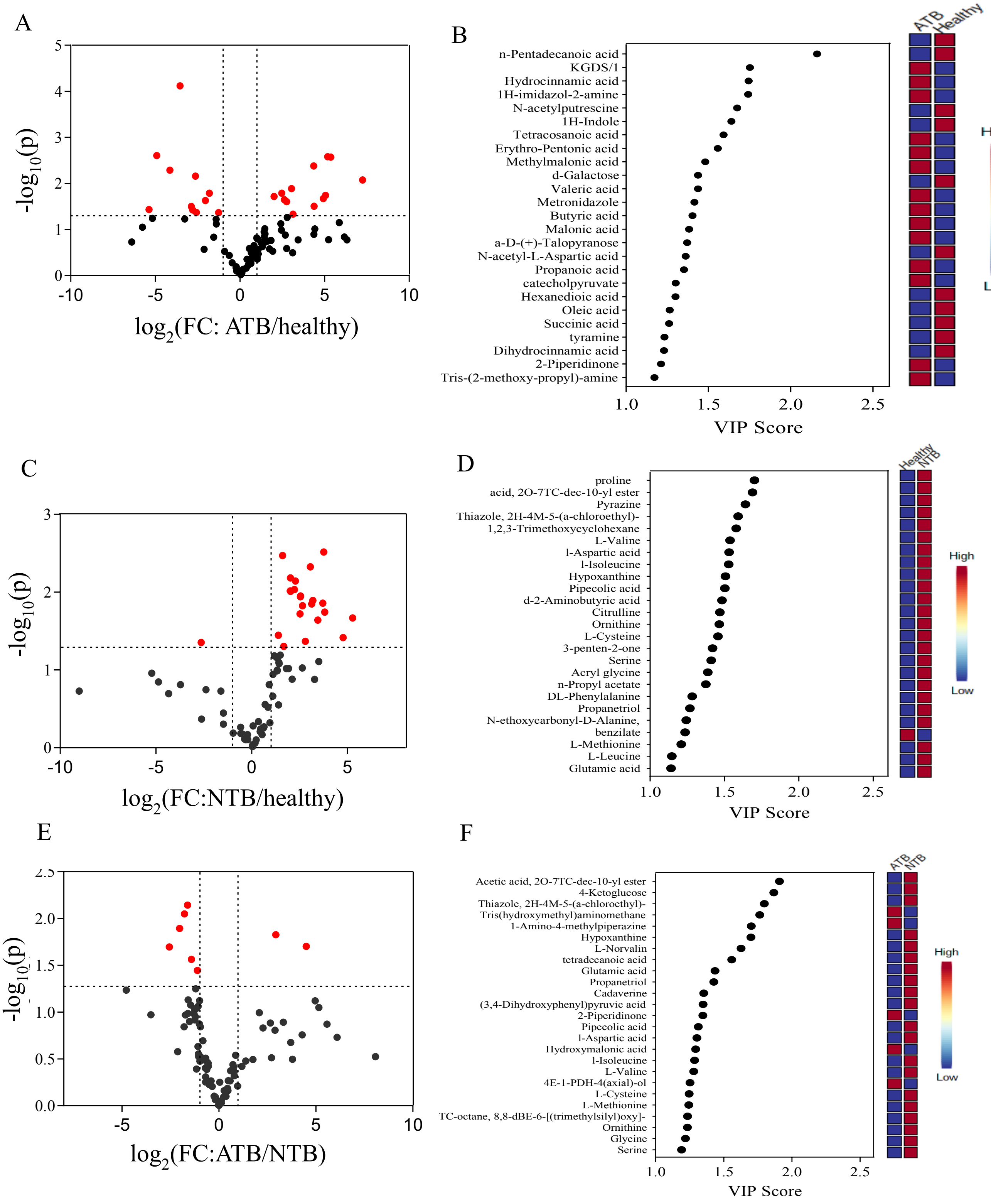


***Figure S6*** *Fecal metabolite analysis of study groups to identify important deregulated molecules from site-I. Volcano plots of A) ATB vs healthy (low/high: 11/13) C) NTB vs healthy (low/high: 8/14) E) ATB vs NTB (low/high: 11/3). Red dots indicate features that presented both a FC ≥ 2 and p-value ≤ 0.05. Important fecal metabolites (the top 25) qualifying VIP score >1 between B) ATB vs healthy D) NTB vs healthy and F) ATB vs NTB. Coloured boxes on the right indicate the relative concentrations of the corresponding metabolite in each study group. N-(4-Bromo-2-fluoro-phenyl)-N'-(3-morpholin-4-yl-propyl)-oxalamide: N-4-BF-N'-3MP-oxalamide.*

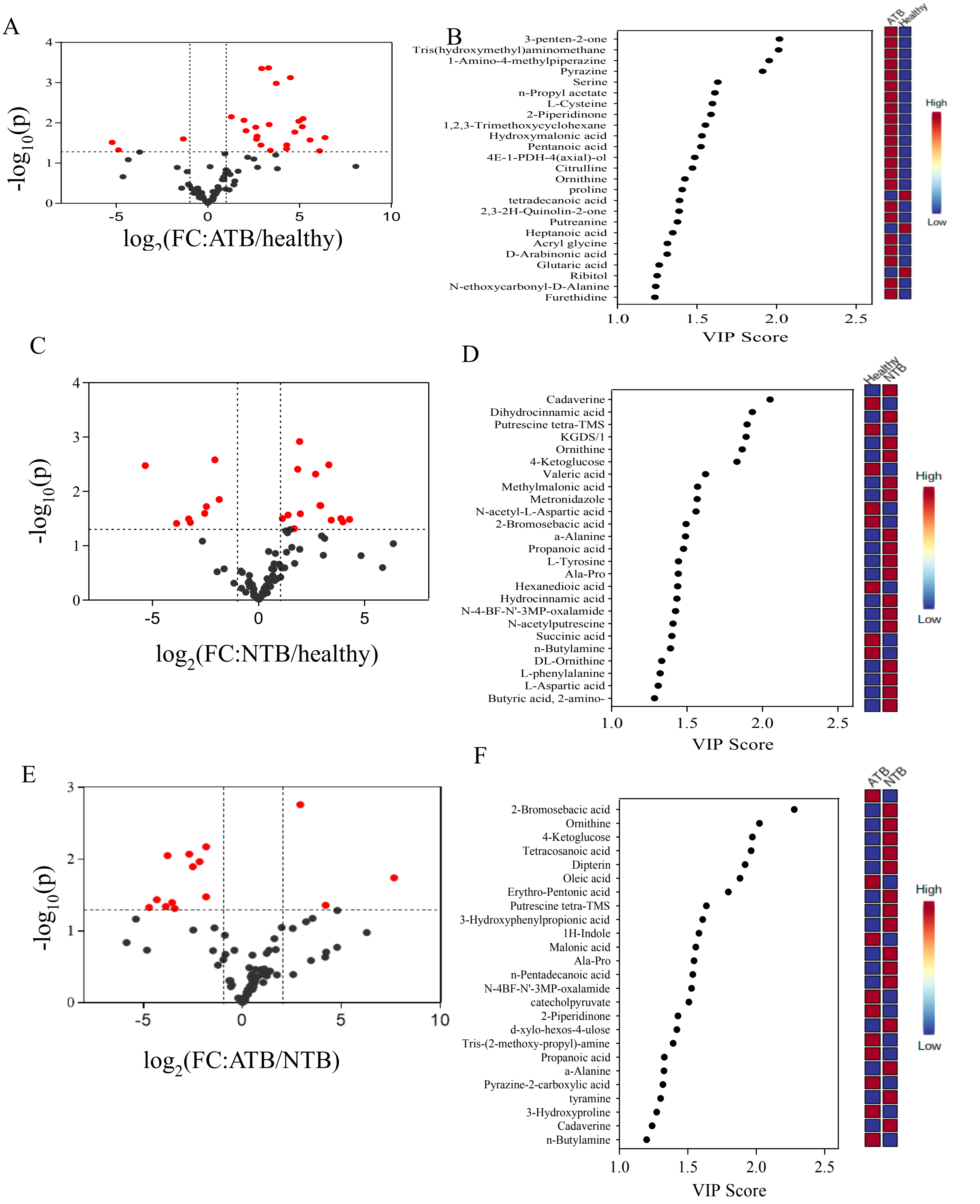


***Figure S7** Fecal metabolite analysis of active tuberculosis patients (ATB), non-tuberculosis (NTB) and healthy subjects of site-II. Volcano plots of A) ATB vs healthy (low/high: 3/22) C) NTB vs healthy (low/high: 1/22) E) ATB vs NTB (low/high: 6/2). Red dots indicate features that presented both a FC ≥ 2 and p-value ≤ 0.05. Rank of the different metabolites (the top 25) identified by the PLS-DA between B) ATB vs healthy D) NTB vs healthy F) ATB vs NTB, according to the VIP score on the x-axis. Coloured boxes on the right indicate the relative concentrations of the corresponding metabolite in each group. Thiazole, 2H-4M-5-(a-chloroethyl)-: Thiazole, 2-hydroxy-4-methyl-5-(a-chloroethyl)-; Acetic acid, 2O-7TC-dec-10-yl ester-: Acetic acid, 2-oxa-7-thia-tricyclo[4.3.1.0(3,8)]dec-10-ylester;TC-octane,8,8-dBE-6-[(trimethylsilyl)oxy]-:*

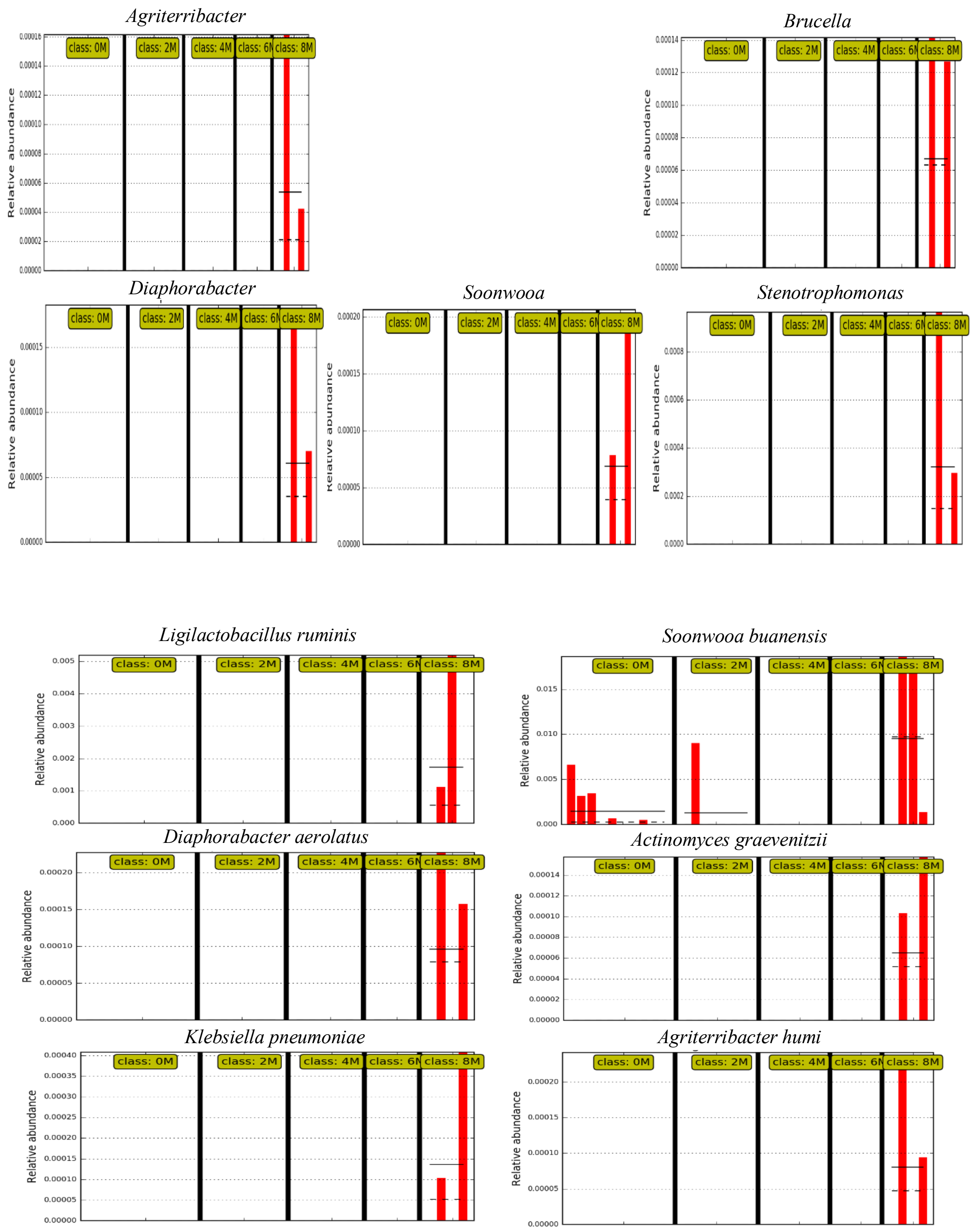


***Figure S8*** *LEfSe analysis of drug-treated followed-up tuberculosis patient samples revealed differential species abundance. Histogram of the relative abundances of genus and species in 0, 2, 4, 6, and 8 month subjects. The mean and median relative abundance of the species are indicated with solid and dashed lines, respectively.*

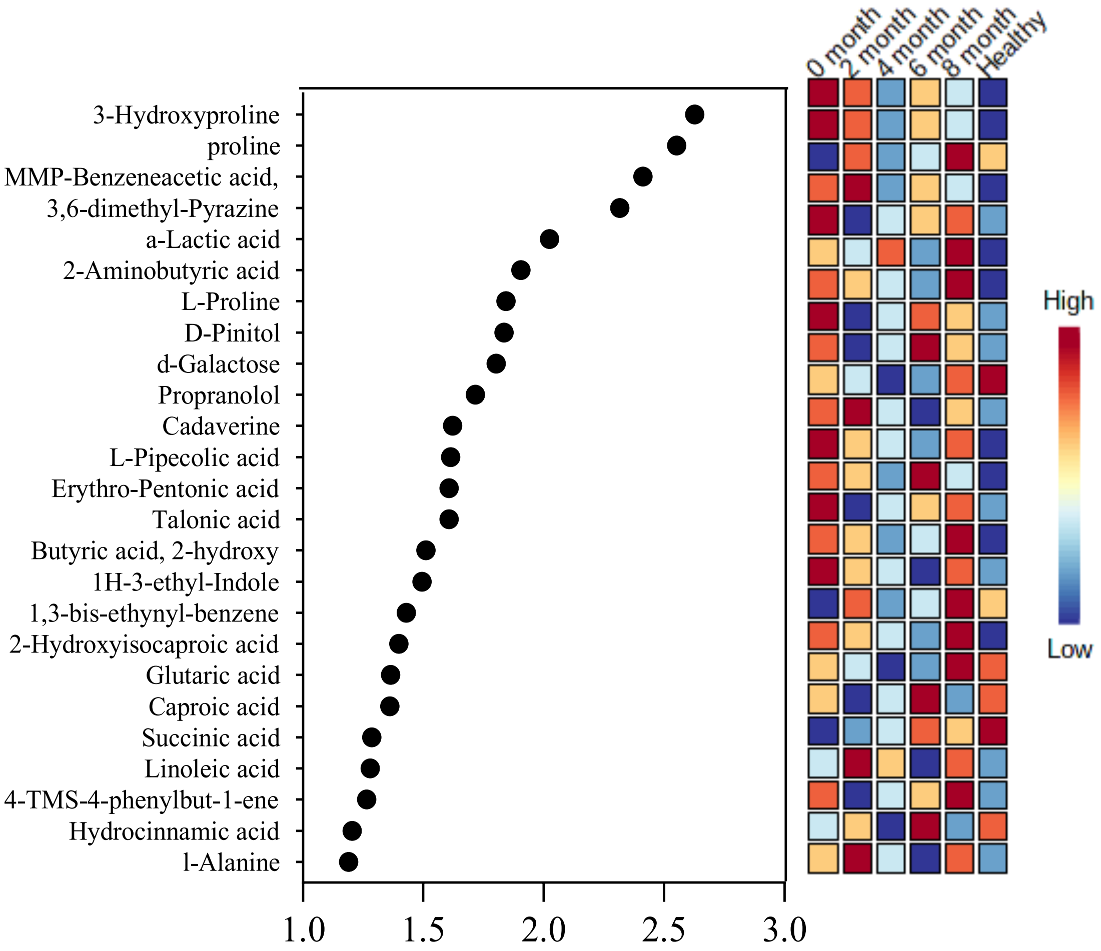


***Figure S9*** *Multivariate analysis of fecal metabolome profile of followed up tuberculosis patients at different months of completing treatment (0/2/4/6/8) and healthy samples using Metaboanalyst 5.0. Rank of the different metabolites (the top 25) identified by the PLS-DA. Coloured boxes on the right indicate the relative concentrations of the corresponding metabolite in each group. a-methyl-4-(2-methylpropyl)-Benzeneacetic acid: MMP-Benzeneacetic acid; 1H-Indole, 1-(trimethylsilyl)-3-[2-[(trimethylsilyl)oxy]ethyl]-: 1H-3-ethyl-Indole ; 4-Trimethylsilyloxy-4-phenylbut-1-ene: 4-TMS-4-phenylbut-1-ene.*

### Supplemental Experimental Procedures

#### DNA extraction and 16SrRNA gene amplicon sequencing

Fecal samples (200 mg) were taken in a pre-chilled microcentrifuge tube (MCT, 1.5 ml). The samples were vortexed after adding TE buffer (200 µl, 50 mM Tris-1mM EDTA, pH 8.0) and glass beads (4 in number, 2.7 mm, Biospec). Lysozyme (50 µl, 10 mg/ml), mutanolysin (6 µl, 2 KU/ml), and lysostaphin (3 µl, 4 KU/ml) were added to the sample and incubated at 37˚ C for 1 hour (hr). To this reaction mixture, guanidinium thiocyanate (250 µl, 4M) was added and mixed for 45 sec and subsequently incubated at 37˚ C for 10 minutes (min) at 300 rpm in Thermomixer C (Eppendorf) after mixing with N-lauryl sarcosine (300 µl of 10 %).

After vortexing and a short spin of 16,000 g for a few seconds, samples were incubated at 70˚C for 1 hr. Then, zirconia beads (300 mg of 0.1 mm, Biospec) were added, followed by bead beating in two cycles of 30 seconds (sec) each, with a total program time of 2 min using a bead beater (Mini bead beater 16, Biospec Products Inc. USA). After bead beating, PVPP (15 mg) was added and vortexed, followed by centrifugation at 16,000 g for 3 min. The supernatant was transferred to a fresh MCT, and the pellet was washed twice with 500 µl of Tris (50 mM)-EDTA (20 mM)-NaCl (100 mM)-PVPP (1%), and the resulting supernatants were pooled. To the supernatant, isopropanol (2 ml) was added and incubated at room temperature for 10 min after inverting the tubes for a few min. The reaction mixture was centrifuged at 16,000 g for 10 min to harvest the pellet and dried at room temperature. To each sample, phosphate buffer: potassium acetate (9:1, 1 ml) was added and incubated overnight at -20°C. The thawed samples were centrifuged at 16,000 g for 30 min at 4˚C. The supernatant was transferred to another tube, and RNase A (4 µl,10 mg/ml) was added before incubating at 37˚C for 30 min. Then sodium acetate (50 µl, 3M) was added to each tube, followed by ice-cold ethanol (1 ml, 96 %) and mixed by inverting the tubes for few times before incubating at room temperature for 5 min. These samples were then centrifuged at 16,000 g for 15 min at 4 ˚C, and after discarding the supernatant, the pellet was washed with ice-cold ethanol (70%). The dried pellet was resuspended in Tris (10 mM)-EDTA (1 mM) buffer (pH 8.0, 200 µl), and harvested genomic DNA was quantified using a Qubit 3.0 fluorometer (Invitrogen, USA).

Extracted genomic DNA was sent to Macrogen, South Korea, for library preparation and sequencing. The libraries were prepared following the Illumina 16S Metagenomic Sequencing Library protocols after amplifying the V3-V4 region of the 16S rRNA gene. The input gDNA (2 ng) was Polymerase Chain Reaction (PCR) amplified with 5X reaction buffer, dNTP mix (1 mM), the universal forward and reverse PCR primer (500 nM each), and Herculase II fusion

DNA polymerase (Agilent Technologies, Santa Clara, CA). The cycle condition for 1st PCR was 3 min at 95 °C for heat activation, and 25 cycles of 30 sec at 95 °C, 30 sec at 55 °C, and 30 sec at 72 °C, followed by a 5 min final extension at 72 °C. The universal primer pair with Illumina adapter overhang sequences used for the first amplification were as follows: V3-F: 5'-TCGTCGGCAGCGTCAGATGTGTATAAGAGACAGCCTACGGGNGGCWGCAG-3', V4-R: 5'- GTCTCGTGGGCTCGGAGATGTGTATAAGAGACAGGACTACHVGGGTATCTAATCC-3. The 1st PCR product was purified with AMPure beads (Agencourt Bioscience, Beverly, MA). Following purification, the 1st PCR product (2 μl) was amplified for final library construction containing the index using NexteraXT Indexed Primer. The cycle condition for the 2nd PCR was the same as the 1st, except it was expanded for 10 cycles. The PCR products were purified with AMPure beads. The final purified products were then quantified using qPCR following the qPCR Quantification Protocol Guide (KAPA Library Quantification kits for Illumina Sequencing platforms) and qualified using the TapeStation D1000 ScreenTape (Agilent Technologies, Waldbronn, Germany).

**Derivatization of fecal metabolites**

The fecal samples (n=94) were randomized before processing, and data acquisition was carried out in multiple batches of 8-12 samples per batch. To protect carbonyl groups and reduce the number of tautomeric isomers, methoxyamine (20 μl) in pyridine (30 mg/ml) was added to the dried fecal metabolites, followed by vortexing for 30 sec and incubation at 37 °C with generous shaking (1000 rpm) for 90 min. The sample vials were inverted once to capture any solvent condensation at the cap surface, followed by a short spin. To derivatize hydroxyl and amine groups of the extracted fecal metabolites to trimethylsilylated (TMS) forms, N-methyl-N-(trimethylsilyl) trifluoroacetamide (MSTFA; 80 μl) with trimethylchlorosilane (1%, TMCS) were added, followed by vortexing for 10 sec and incubation at 37 °C with shaking (1000 rpm) for 30 min. After derivatization, samples were centrifuged at 10,000 g for 5 min, and the supernatant was transferred to a 200 μl glass vial insert in a 2 ml vial. The derivatized fecal metabolites were brought to room temperature, and mass spectrometry data acquisition was carried out within 24 h of derivatization. An equal volume of extracted fecal metabolites of all samples was pooled to prepare a quality control (QC) sample. Aliquots (300 μl each) of the QC were used for derivatization using the same procedure, and at least three QC samples were run in each batch.
